# Associations between motor and executive/attentional deficits in cerebral palsy: Scoping review

**DOI:** 10.1101/2025.01.26.25321141

**Authors:** Melinda Martinet, Kathrin Reichert, Juliane Schneider, Dajie Zhang, Peter B. Marschik, Pawel J. Matusz

## Abstract

**Background:** Cerebral palsy (CP) is the leading cause of motor disability in children and is frequently accompanied by cognitive impairments, including deficits in executive and attentional functions. Despite growing research on motor-cognitive interactions in other populations, evidence in CP remains limited. Objective: To identify gaps in the current literature, this review synthesised evidence on the relationships between specific motor and executive/attentional functions in CP.

**Method:** A systematic search of three databases was conducted. Articles were screened using predefined eligibility criteria by two independent assessors. The results were summarised in tables and narrative form. The review was written in accordance with the JBI Guidelines.

**Results:** Thirteen articles were included in the review, spanning 652 participants aged between 22 months and 62 years. Five studies reported significant links of motor functions with executive functions, and five with attentional functions. Gross and fine motor skills were most reliably linked to working memory, fine motor skills to overall executive functioning, and gross motor skills to focused attention. Three studies found no evidence of links between motor and cognitive control domains. We include an overview of tools used for measuring motor and attention/executive functions in the reviewed studies.

**Conclusions:** Our review suggests positive associations between motor and executive/attentional functions in individuals with CP, highlighting their interdependence. More longitudinal empirical studies are needed, to confirm these links, which will help design better interventions. Despite the still limited evidence, our findings and anecdotal clinical reports call for intervention studies evaluating the multidimensional, cross-domain functional improvements.

## Introduction

Cerebral palsy (CP) is the leading cause of motor disability in children (Paul et al., 2022), with a global prevalence of 1.6 per 1,000 births (McIntyre et al., 2022). CP is defined as “a group of permanent disorders of the development of movement and posture, causing activity limitation, that are attributed to non-progressive disturbances that occurred in the developing fetal or infant brain” (Rosenbaum et al., 2006, p.9). CP is diagnosed and classified by the presence of specific disorders of movement (e.g., motor control, coordination, posture) or muscle tone, in lower/upper limbs, on one or both sides of the body (Rosenbaum et al. 2006; Paul et al., 2022). Although the underlying brain disturbance is non-progressive, its clinical manifestations may change across the lifespan. Consequently, the primary motor disorders may with time lead to the development of secondary musculoskeletal problems, jointly limiting activity (Paul et al., 2022; Rosenbaum et al., 2007). The degree of independent motor functioning vary widely in CP (Rosenbaum et al. 2026, Paul et al., 2022; Vitrikas et al., 2020). Gross motor functioning is evaluated by the severity of the underlying motor disorders via standard systems, such as, e.g., the Gross Motor Function Classification System (GMFCS; Palisano et al. 2008), which ranges from largely independent mobility (Level I) to severe limitations in voluntary movement and self-mobility (Level V). Gross motor difficulties may involve postural control, balance and locomotion, but upper-limb and fine-motor functions may also be affected, including reaching, grasping or writing (Paul et al., 2022; Van Eck et al., 2009).

CP is also frequently accompanied by abnormalities in mental functioning and physical health (Rosenbaum et al., 2007; Vitrikas et al., 2020). Mental functioning in CP likewise varies considerably, from severe impairment to typical-range sensation-perception, cognition, and communication (Stadskleiv et al., 2018; Wotherspoon et al., 2023). Among cognitive domains, attentional and executive functions are particularly relevant to motor behaviour, as goal-directed action requires the selection of relevant information and flexible adaptation to changing environmental demands (Diamond 2000, Miyake et al. 2000). Attention is an umbrella term referring to the cognitive processes that prioritise and select information relevant to current goals (Lodha & Gupta, 2022). Five types of attention are often distinguished: focused attention (ability to respond to specific stimuli), sustained attention (maintenance of attention across time), selective attention (prioritisation of relevant information despite distraction), alternating attention (shifting of attention between tasks) and divided attention (allocation of attention to multiple concurrent demands; Sohlberg & Mateer, 2001). The most often studied executive functions are working memory (maintenance of information in an active state in one’s mind), cognitive flexibility (goal-based adaptation to changing demands) and inhibition (suppressing the dominating but incorrect responses).

In CP, difficulties are common in both motor and cognitive control (attentional and executive) functions. Substantial attention or executive-function difficulties are reported in ∼30–50% of children with CP, while attention-deficit/hyperactivity disorder (ADHD) is diagnosed in around one-fifth to one-third of them (e.g. Casseus et al., 2024; Kasap et al., 2024; Maioli et al., 2019; Pirila et al., 2011). Children with CP have more issues with sustained and divided attention, inhibition, working memory and cognitive flexibility (e.g. Bottcher et al., 2010; Cabezas & Carriedo, 2020). Additionally, there is growing evidence that difficulties in attention and executive functions also contribute to limitations in everyday activities and participation, suggesting their functional importance (Bottcher, 2010; Mousavi et al., 2023). However, it currently remains unclear if motor difficulties are reliably linked to attentional and/or executive impairments found in CP. It is important to verify if such relationships exist, both to better characterize and understand CP as a disorder, but also to offer important insights into care for individuals with CP.

There is strong converging evidence that the development of motor and cognitive control functions is inter-connected. In the context of early identification of CP, the assessment of spontaneous motor function, known general movements (GMs), has proven to be a sensitive marker of healthy neurodevelopment (e.g., Prechtl et al 1997; Novak et al 2018). Emerging during the prenatal period (Einspieler et al 2021), GMs scaffold the development of more complex motor functions (e.g., Einspieler et al 2008) as well as cognitive functions, including language and cognitive capacities (e.g., Olsen et al. 2018; Einspieler et al. 2016; Salavati et al. 2017). Likewise, difficulties in postural control delays early language and attention development (e.g., Dusing & Harboune, 2010). Motor skills have also been predict long-term executive functions, in both typically developing children and those with neurodevelopmental impairments at school-age (e.g., Bernardi et al., 2018; Piek et al., 2008; Zhou & Tolmie, 2024). Motor and attentional/executive skills continue to show consistent relationships throughout childhood and adolescence, at both general and process-specific levels (Gandotra et al., 2021; Bao et al. 2024). This inter-dependence may be explained by several mechanisms. Both functions undergo important development between the age of 5 and 10 years, offering ample opportunities for interaction (Gabbard, 2008). They also both rely on common processes, like sequencing, monitoring and planning, and both engage the prefrontal cortex and cerebellum (e.g., Diamond et al. 2000; Sergeant, 2000). Notably, these associations are even stronger in children with developmental disorders and delays. These strong associations are observed in both nominally “cognitive” (ADHD) and “motor” conditions (developmental coordination disorder; e.g., Houwen et al. 2016; Klupp et al. 2021; Fogel et al. 2021).

Understanding the presence and strength of the cognitive control–motor relationships in CP is important for better understanding of this condition. According to Thelen and Smith, developmental abilities emerge from reciprocal, non-linear interactions among neural, bodily, task and environmental constraints. As a result, atypicalities in motor capacities change how children explore and act on their environment and so may modify the opportunities and demands through which children’s attentional and executive functions develop (Thelen & Smith, 1994; Smith & Thelen, 2003). Similarly, Hadders-Algra proposes that development involves an initially broad repertoire of neural and behavioural solutions, followed by experience-dependent selection of increasingly adaptive strategies. The early brain injury in CP may restrict both this repertoire and the capacity for adaptive strategies, potentially altering developmental trajectories beyond the immediately affected motor behaviour, to how attention is controlled (Hadders-Algra, 2000, 2010). However, co-occurring motor and cognitive-control difficulties in CP do not necessarily imply that impairments in the two domains are developmentally coupled. The extent of brain injury has been reported to predict the severity of both motor and cognitive difficulties (Rai et al., 2013; Reid et al. 2015). Thus, the co-occurrence of the latter could represent at least partly independent consequences of the same early-life brain injury. Evidence from the rare existing intervention studies likewise provides only partial support for links between the motor and cognitive control domains. Recent reviews examining physical and physical-cognitive interventions in CP report overall consistent benefits for attention, but more variable effects in executive functions (Blasco et al. 2023; Carracedo-Martín et al., 2025). Hence, it remains unclear not only if CP is characterized by a general relationship between motor and cognitive-control functioning, but also if associations between particular motor attentional/executive processes are reliable.

Establishing the nature of these relationships has direct clinical implications. If the severity of motor difficulties reliably signals attentional or executive impairment, motor assessments could help identify children requiring more comprehensive cognitive phenotyping and support integrated treatment targeting the broader functional profile of CP (e.g. Marschik et al. 2017). Additionally, this indicates that physical activity or exergames, which are cost-effective and easy to integrate in everyday life interventions, could have important wide-range benefits for CP. Such interventions indeed improve executive and attentional functions in typical and ADHD populations (Benzing et al., 2018; de Greeff et al., 2018; Zhu et al., 2023). Conversely, if the motor– cognitive control associations are weak, inconsistent or limited to particular skill pairings, motor and cognitive-control domains would need to be assessed, and potentially treated, independently in each child. The present review therefore examined whether, across existing observational studies, the level of motor impairment in children and adolescents with CP is associated with the level of attentional and/or executive-function impairment, and whether any such associations are domain-general or specific to particular combinations of functions.

## Method

This review was conducted and reported in accordance with the Joanna Briggs Institute (JBI) manual for evidence synthesis (Peters et al., 2020).

### Search strategy

To formulate the research equation, three key concepts were identified: (1) cerebral palsy, (2) motor functions and (3) executive and attentional functions. In order to determine the relevant key terms for each concept, an analysis of the Babik et al. (2023) article, identified during the preliminary research, as well as its literary references, was carried out. The full list of key terms is given in **Error! Reference source not found**..

Searches for this review were conducted on three databases: PubMed, Embase and Cochrane. The initial search equation yielded a broad selection of articles on these three platforms. By analysing the results and collaborating with faculty librarians, the search parameters were refined to develop the final search equation [Appendix 1

*Table 2* : *Research equation*

]. A filter was applied to exclude articles written in languages other than English, French or German. The last search was carried out on 14 February 2024 and updated on 15 March 2026.

### Selection process

To identify relevant literary research articles, inclusion and exclusion criteria were established. The criteria were chosen by analysing those used in similar reviews (e.g. Fogel et al., 2023; Gandotra et al., 2021). Inclusion criteria were studies that: (1) included patients with a diagnosis of cerebral palsy; (2) addressed motor functions and executive/attentional functions; (3) assessed the links between motor and executive/attentional functions; (4) were written in English, French or German. Exclusion criteria were studies that: (1) were protocols, conference abstracts, and commentaries; (2) addressed only motor functions or executive/attentional functions but not both; (3) made no link between motor and cognitive measures. Study selection was conducted independently by the first two authors with disagreements at any stage at any stage solved by consensus. The first stage of the selection process involved an evaluation of article titles and abstracts according to the inclusion and exclusion criteria (see below). The subsequent stage involved reading of the full article texts, applying the same inclusion and exclusion criteria.

### Data extraction & summary

One primary author extracted the data for the selected articles. The second primary author then verified the extracted results, and any disagreements were resolved through discussion between the two first authors or with the senior author, in case of a lack of consensus. The data extracted included: (1) title, author, and year of publication, (2) study design, (3) participant characteristics (sample size, age, gender, type of CP, and GMFCS and/or MACS^1^ scores), (4) motor functions measured and their corresponding results, (5) executive and/or attentional functions measured and their corresponding results and finally (6) type, direction and strength of the relationship observed between motor and cognitive functions. The extracted main results are presented in Table 3. The results were grouped by specific age ranges (subdivision) and subsequently organised according to the motor domains assessed (first studies testing both gross and fine motor functions, then gross motor functions, lastly fine motor functions). Within these categories, studies were further ordered by the type of cognitive control function tested (first studies testing both executive and attentional functions, followed by executive functions only, followed by attentional functions). Synthesis of the main findings in the Table 3 was supported by a narrative summary. In addition, Table 2 was created, to describe the relevant motor and cognitive control measures t used in the included articles.

## Results

### Selection of studies

The selection process is illustrated as a flow diagram in **Error! Reference source not found**., in accordance with the PRISMA-ScR (Preferred Reporting Items for Systematic Reviews & Meta-analysis extension for Scoping

Reviews; Tricco et al. 2018). The search on the three databases identified a total of 486 articles. After exclusion of duplicates, 327 articles were retained for the selection process. After screening for title and abstract, 284 articles were discarded. Subsequently, 33 articles were excluded after a full reading of the article texts. At this stage, one article was added. It corresponds to an article in conference proceedings that had been excluded during the earlier selection stages. In March 2026, a new search was launched, and 2 new articles were added. In the end, a total of 13 articles were retained for this review.

Of these selected articles, 12 were cross-sectional studies (Reilly et al. 2008; Forsman & Eliasson, 2016; Soriano & Hustad, 2021; Stadskleiv et al., 2018; Al-Nemr & Abdelazeim, 2018; Van Rooijen et al., 2012, 2016; Ballester-Plané et al., 2018; Wang et al., 2020; Wotherspoon et al., 2023; Kalkantzi et al. 2025; Abd El Rahman & Sayed 2025), and 1 pre-post interventional study (Surkar et al., 2015). 3 presents a description of all the relevant information from the studies selected that met the objectives of this scoping review, in accordance with the PRISMA-ScR (Tricot et al. 2018).

**Figure 1.**
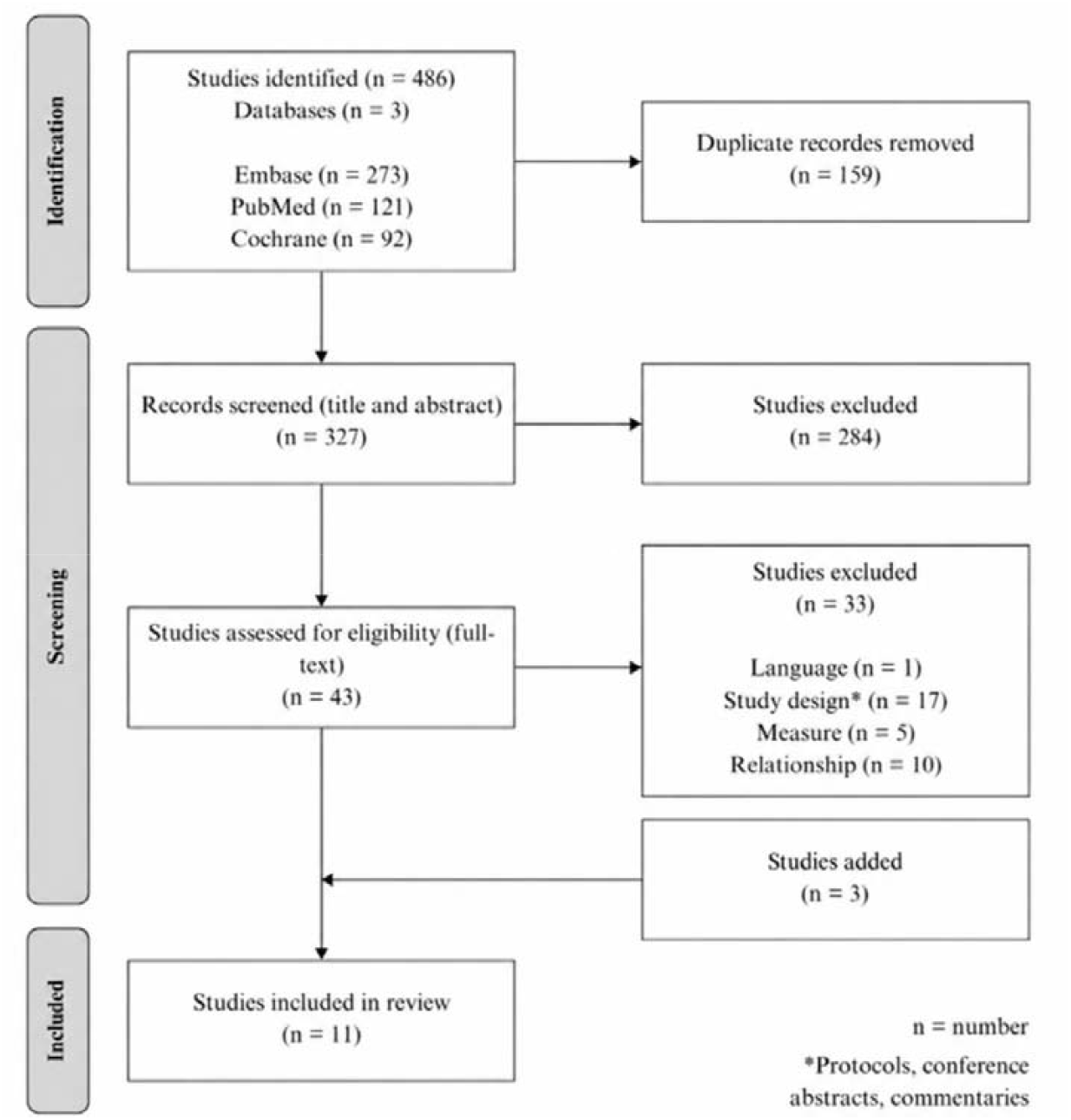
PRISMA-ScR Flow diagram of article selection for this scoping review

### Measures of motor and cognitive control skills

Table 2 describes the tools used in the reviewed studies to measure the motor, and executive and attentional functions, and summarises how many studies used each test (Section *a* and *b*, respectively). Table 3 presents which study used which test for which function. For measures of motor functions, 3 studies assessed both gross and fine motor functions, with a total of 9 studies measuring gross motor function and 6 fine motor function. Most of the studies evaluated general motor abilities, with 4 measuring specific gross motor skills, like postural control, sitting, or walking, and 3 specific fine motor skills, like bimanual function. For measures of cognitive control, 3 articles measured both executive functions and attention, with executive functions reported in a total of 9 studies and attention in 7 studies. Here, 4 studies tested working memory, 3 general executive functioning, 2 cognitive flexibility; 2 tested focused attention specifically, with other single studies testing general and other specific types of attention (e.g., selective, spatial, verbal).

**Table 1.**
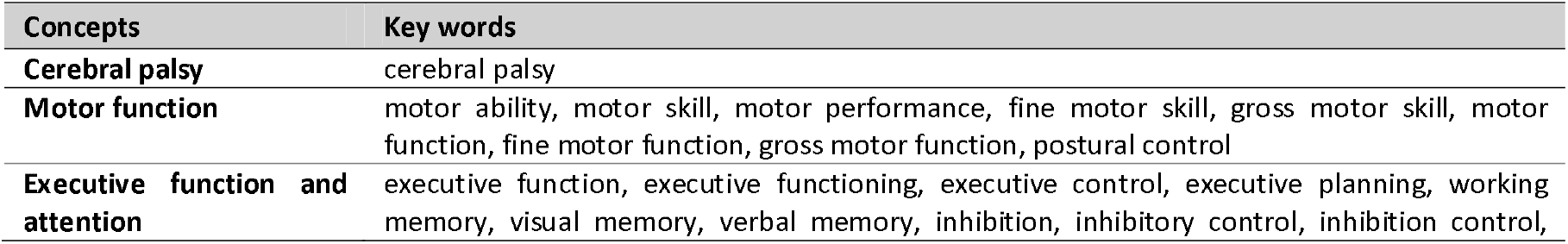
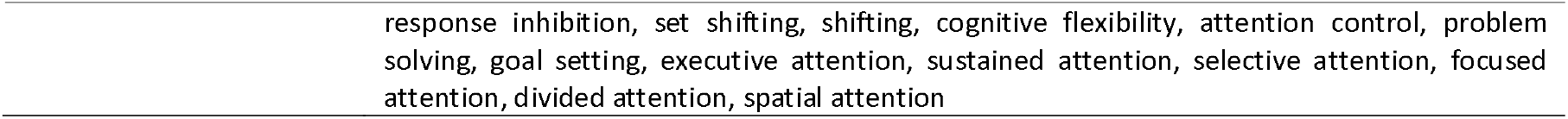
Concepts and keywords for formulating the research equation.

**Table 2.**
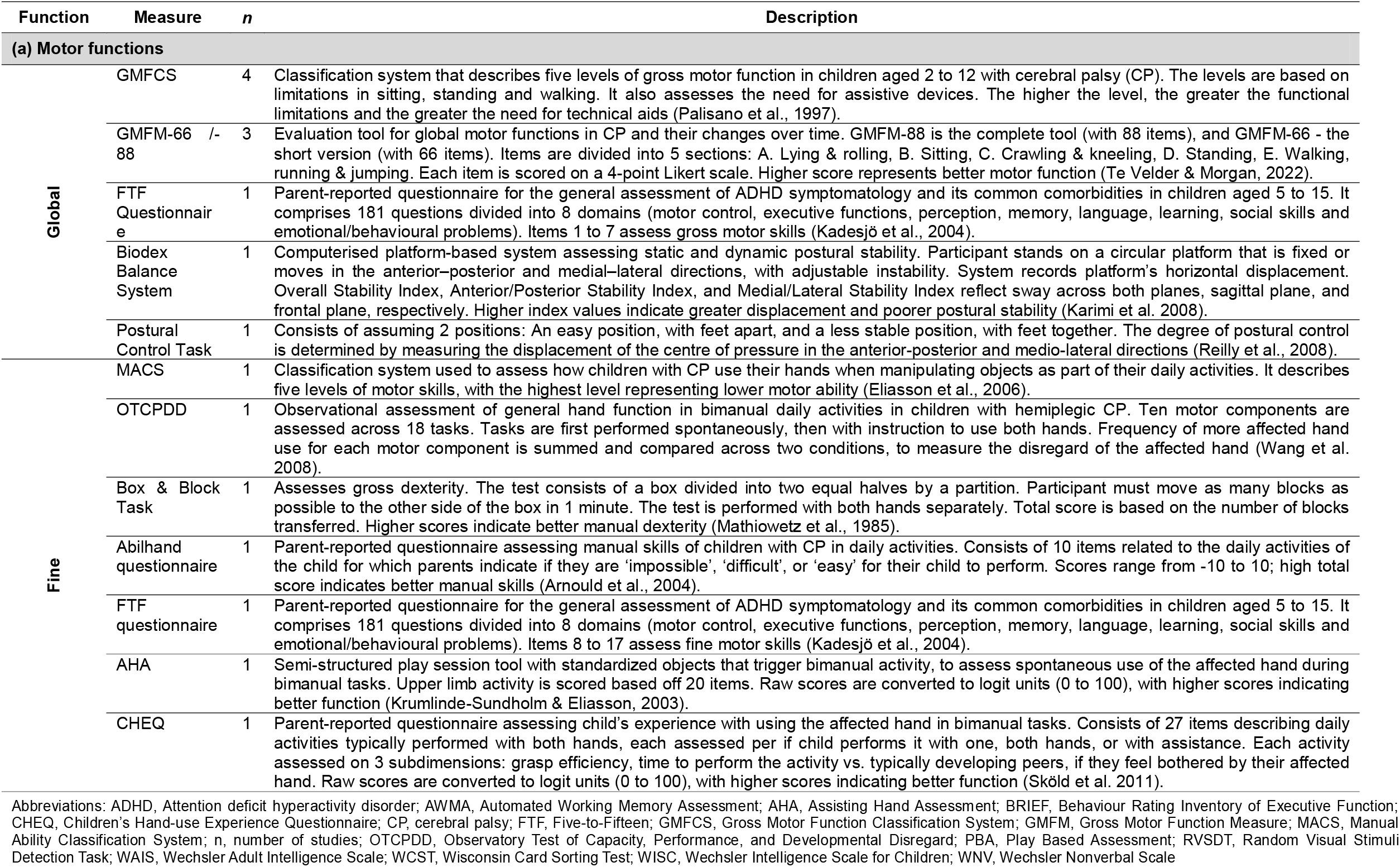
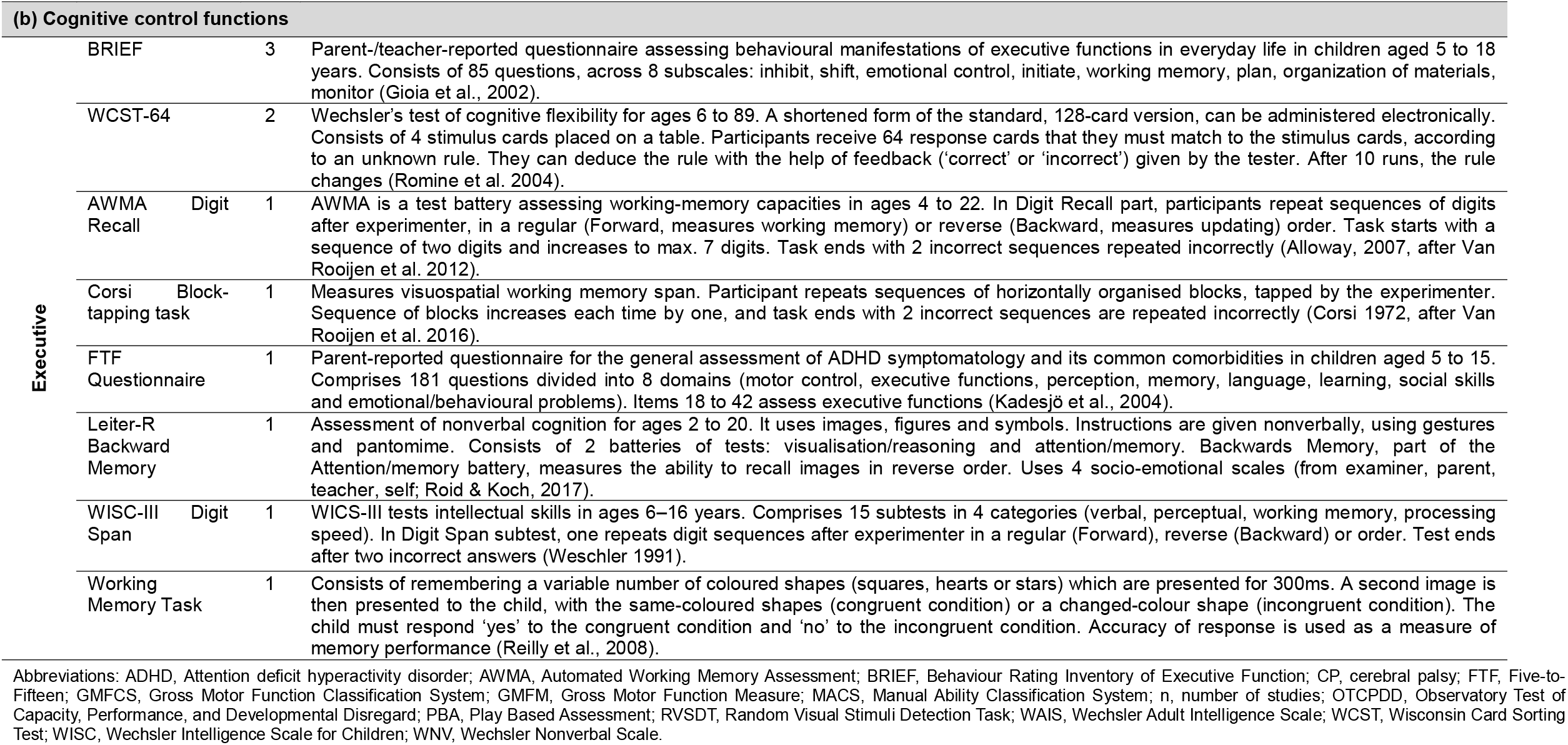
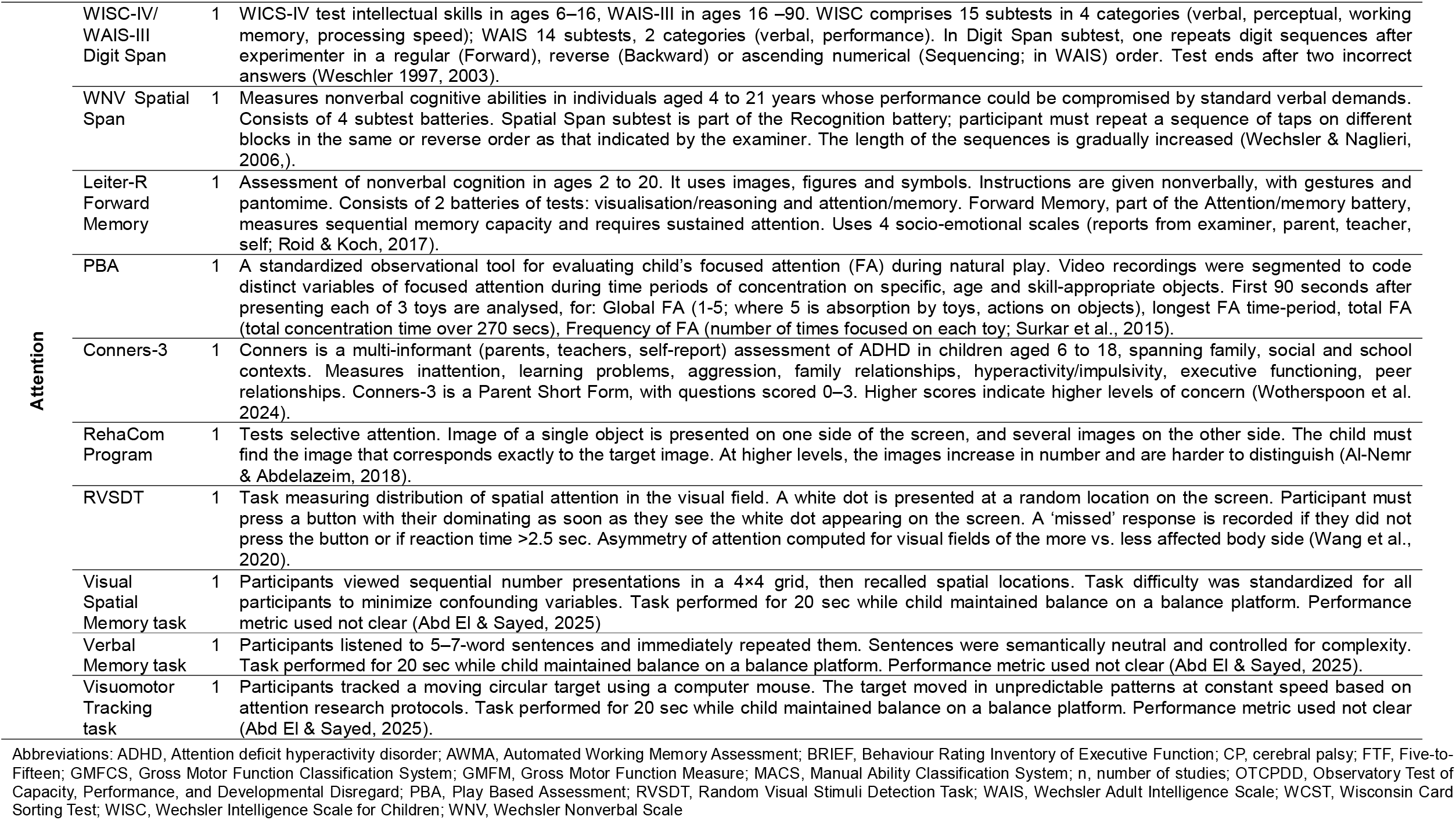
Descriptions of the tests used in the reviewed studies to measure cognitive control-motor links.

| Function | Measure | n | Description |
| --- | --- | --- | --- |
| **(a) Motor functions** |  |  |  |
| **Global** | GMFCS | 4 | Classification system that describes five levels of gross motor function in children aged 2 to 12 with cerebral palsy (CP). The levels are based on limitations in sitting, standing and walking. It also assesses the need for assistive devices. The higher the level, the greater the functional limitations and the greater the need for technical aids (Palisano et al., 1997). |
|  | GMFM-66 /- 88 | 3 | Evaluation tool for global motor functions in CP and their changes over time. GMFM-88 is the complete tool (with 88 items), and GMFM-66 - the short version (with 66 items). Items are divided into 5 sections: A. Lying & rolling, B. Sitting, C. Crawling & kneeling, D. Standing, E. Walking, running & jumping. Each item is scored on a 4-point Likert scale. Higher score represents better motor function (Te Velder & Morgan, 2022). |
|  | FTF Questionnaire | 1 | Parent-reported questionnaire for the general assessment of ADHD symptomatology and its common comorbidities in children aged 5 to 15. It comprises 181 questions divided into 8 domains (motor control, executive functions, perception, memory, language, learning, social skills and emotional/behavioural problems). Items 1 to 7 assess gross motor skills (Kadesjö et al., 2004). |
|  | Biodex Balance System | 1 | Computerised platform-based system assessing static and dynamic postural stability. Participant stands on a circular platform that is fixed or moves in the anterior-posterior and medial-lateral directions, with adjustable instability. System records platform's horizontal displacement. Overall Stability Index, Anterior/Posterior Stability Index, and Medial/Lateral Stability Index reflect sway across both planes, sagittal plane, and frontal plane, respectively. Higher index values indicate greater displacement and poorer postural stability (Karimi et al. 2008). |
|  | Postural Control Task | 1 | Consists of assuming 2 positions: An easy position, with feet apart, and a less stable position, with feet together. The degree of postural control is determined by measuring the displacement of the centre of pressure in the anterior-posterior and medio-lateral directions (Reilly et al., 2008). |
| **Fine** | MACS | 1 | Classification system used to assess how children with CP use their hands when manipulating objects as part of their daily activities. It describes five levels of motor skills, with the highest level representing lower motor ability (Eliasson et al., 2006). |
|  | OTCPDD | 1 | Observational assessment of general hand function in bimanual daily activities in children with hemiplegic CP. Ten motor components are assessed across 18 tasks. Tasks are first performed spontaneously, then with instruction to use both hands. Frequency of more affected hand use for each motor component is summed and compared across two conditions, to measure the disregard of the affected hand (Wang et al. 2008). |
|  | Box & Block Task | 1 | Assesses gross dexterity. The test consists of a box divided into two equal halves by a partition. Participant must move as many blocks as possible to the other side of the box in 1 minute. The test is performed with both hands separately. Total score is based on the number of blocks transferred. Higher scores indicate better manual dexterity (Mathiowetz et al., 1985). |
|  | Abilhand questionnaire | 1 | Parent-reported questionnaire assessing manual skills of children with CP in daily activities. Consists of 10 items related to the daily activities of the child for which parents indicate if they are 'impossible', 'difficult', or 'easy' for their child to perform. Scores range from -10 to 10; high total score indicates better manual skills (Arnould et al., 2004). |
|  | FTF questionnaire | 1 | Parent-reported questionnaire for the general assessment of ADHD symptomatology and its common comorbidities in children aged 5 to 15. It comprises 181 questions divided into 8 domains (motor control, executive functions, perception, memory, language, learning, social skills and emotional/behavioural problems). Items 8 to 17 assess fine motor skills (Kadesjö et al., 2004). |
|  | AHA | 1 | Semi-structured play session tool with standardized objects that trigger bimanual activity, to assess spontaneous use of the affected hand during bimanual tasks. Upper limb activity is scored based off 20 items. Raw scores are converted to logit units (0 to 100), with higher scores indicating better function (Krumlinde-Sundholm & Eliasson, 2003). |
|  | CHEQ | 1 | Parent-reported questionnaire assessing child's experience with using the affected hand in bimanual tasks. Consists of 27 items describing daily activities typically performed with both hands, each assessed per if child performs it with one, both hands, or with assistance. Each activity assessed on 3 subdimensions: grasp efficiency, time to perform the activity vs. typically developing peers, if they feel bothered by their affected hand. Raw scores are converted to logit units (0 to 100), with higher scores indicating better function (Sköld et al. 2011). |
| **(b) Cognitive control functions** |  |  |  |
| Executive | BRIEF | 3 | Parent-/teacher-reported questionnaire assessing behavioural manifestations of executive functions in everyday life in children aged 5 to 18 years. Consists of 85 questions, across 8 subscales: inhibit, shift, emotional control, initiate, working memory, plan, organization of materials, monitor (Gioia et al., 2002). |
|  | WCST-64 | 2 | Wechsler's test of cognitive flexibility for ages 6 to 89. A shortened form of the standard, 128-card version, can be administered electronically. Consists of 4 stimulus cards placed on a table. Participants receive 64 response cards that they must match to the stimulus cards, according to an unknown rule. They can deduce the rule with the help of feedback ('correct' or 'incorrect') given by the tester. After 10 runs, the rule changes (Romine et al. 2004). |
|  | AWMA<br>Digit Recall | 1 | AWMA is a test battery assessing working-memory capacities in ages 4 to 22. In Digit Recall part, participants repeat sequences of digits after experimenter, in a regular (Forward, measures working memory) or reverse (Backward, measures updating) order. Task starts with a sequence of two digits and increases to max. 7 digits. Task ends with 2 incorrect sequences repeated incorrectly (Alloway, 2007, after Van Rooijen et al. 2012). |
|  | Corsi<br>Block-tapping task | 1 | Measures visuospatial working memory span. Participant repeats sequences of horizontally organised blocks, tapped by the experimenter. Sequence of blocks increases each time by one, and task ends with 2 incorrect sequences are repeated incorrectly (Corsi 1972, after Van Rooijen et al. 2016). |
|  | FTF<br>Questionnaire | 1 | Parent-reported questionnaire for the general assessment of ADHD symptomatology and its common comorbidities in children aged 5 to 15. Comprises 181 questions divided into 8 domains (motor control, executive functions, perception, memory, language, learning, social skills and emotional/behavioural problems). Items 18 to 42 assess executive functions (Kadesjö et al., 2004). |
|  | Leiter-R<br>Backward Memory | 1 | Assessment of nonverbal cognition for ages 2 to 20. It uses images, figures and symbols. Instructions are given nonverbally, using gestures and pantomime. Consists of 2 batteries of tests: visualisation/reasoning and attention/memory. Backwards Memory, part of the Attention/memory battery, measures the ability to recall images in reverse order. Uses 4 socio-emotional scales (from examiner, parent, teacher, self; Roid & Koch, 2017). |
|  | WISC-III<br>Digit Span | 1 | WISC-III tests intellectual skills in ages 6–16 years. Comprises 15 subtests in 4 categories (verbal, perceptual, working memory, processing speed). In Digit Span subtest, one repeats digit sequences after experimenter in a regular (Forward), reverse (Backward) or order. Test ends after two incorrect answers (Wechsler 1991). |
|  | Working<br>Memory Task | 1 | Consists of remembering a variable number of coloured shapes (squares, hearts or stars) which are presented for 300ms. A second image is then presented to the child, with the same-coloured shapes (congruent condition) or a changed-colour shape (incongruent condition). The child must respond 'yes' to the congruent condition and 'no' to the incongruent condition. Accuracy of response is used as a measure of memory performance (Reilly et al., 2008). |
| **(b) Cognitive control functions: continued** |  |  |  |
| Attention | WISC-IV/<br>WAIS-III<br>Digit Span | 1 | WISC-IV test intellectual skills in ages 6–16, WAIS-III in ages 16–90. WISC comprises 15 subtests in 4 categories (verbal, perceptual, working memory, processing speed); WAIS 14 subtests, 2 categories (verbal, performance). In Digit Span subtest, one repeats digit sequences after experimenter in a regular (Forward), reverse (Backward) or ascending numerical (Sequencing; in WAIS) order. Test ends after two incorrect answers (Wechsler 1997, 2003). |
|  | WNV Spatial Span | 1 | Measures nonverbal cognitive abilities in individuals aged 4 to 21 years whose performance could be compromised by standard verbal demands. Consists of 4 subtest batteries. Spatial Span subtest is part of the Recognition battery; participant must repeat a sequence of taps on different blocks in the same or reverse order as that indicated by the examiner. The length of the sequences is gradually increased (Wechsler & Naglieri, 2006.). |
|  | Leiter-R<br>Forward<br>Memory | 1 | Assessment of nonverbal cognition in ages 2 to 20. It uses images, figures and symbols. Instructions are given nonverbally, with gestures and pantomime. Consists of 2 batteries of tests: visualisation/reasoning and attention/memory. Forward Memory, part of the Attention/memory battery, measures sequential memory capacity and requires sustained attention. Uses 4 socio-emotional scales (reports from examiner, parent, teacher, self; Roid & Koch, 2017). |
|  | PBA | 1 | A standardized observational tool for evaluating child's focused attention (FA) during natural play. Video recordings were segmented to code distinct variables of focused attention during time periods of concentration on specific, age and skill-appropriate objects. First 90 seconds after presenting each of 3 toys are analysed, for: Global FA (1-5; where 5 is absorption by toys, actions on objects), longest FA time-period, total FA (total concentration time over 270 secs), Frequency of FA (number of times focused on each toy; Surkar et al., 2015). |
|  | Conners-3 | 1 | Conners is a multi-informant (parents, teachers, self-report) assessment of ADHD in children aged 6 to 18, spanning family, social and school contexts. Measures inattention, learning problems, aggression, family relationships, hyperactivity/impulsivity, executive functioning, peer relationships. Conners-3 is a Parent Short Form, with questions scored 0–3. Higher scores indicate higher levels of concern (Wotherspoon et al. 2024). |
|  | RehaCom<br>Program | 1 | Tests selective attention. Image of a single object is presented on one side of the screen, and several images on the other side. The child must find the image that corresponds exactly to the target image. At higher levels, the images increase in number and are harder to distinguish (Al-Nemr & Abdelazeim, 2018). |
|  | RVSDT | 1 | Task measuring distribution of spatial attention in the visual field. A white dot is presented at a random location on the screen. Participant must press a button with their dominating as soon as they see the white dot appearing on the screen. A 'missed' response is recorded if they did not press the button or if reaction time >2.5 sec. Asymmetry of attention computed for visual fields of the more vs. less affected body side (Wang et al., 2020). |
|  | Visual<br>Spatial<br>Memory task | 1 | Participants viewed sequential number presentations in a 4×4 grid, then recalled spatial locations. Task difficulty was standardized for all participants to minimize confounding variables. Task performed for 20 sec while child maintained balance on a balance platform. Performance metric used not clear (Abd El & Sayed, 2025) |
|  | Verbal<br>Memory task | 1 | Participants listened to 5–7-word sentences and immediately repeated them. Sentences were semantically neutral and controlled for complexity. Task performed for 20 sec while child maintained balance on a balance platform. Performance metric used not clear (Abd El & Sayed, 2025). |
|  | Visuomotor<br>Tracking<br>task | 1 | Participants tracked a moving circular target using a computer mouse. The target moved in unpredictable patterns at constant speed based on attention research protocols. Task performed for 20 sec while child maintained balance on a balance platform. Performance metric used not clear (Abd El & Sayed, 2025). |

Abbreviations: ADHD, Attention deficit hyperactivity disorder; AWMA, Automated Working Memory Assessment; BRIEF, Behaviour Rating Inventory of Executive Function; CP, cerebral palsy; FTF, Five-to-Fifteen; GMFCS, Gross Motor Function Classification System; GMFM, Gross Motor Function Measure; MACS, Manual Ability Classification System; n, number of studies; OTCPDD, Observatory Test of Capacity, Performance, and Developmental Disregard; PBA, Play Based Assessment; RVSDT, Random Visual Stimuli Detection Task; WAIS, Wechsler Adult Intelligence Scale; WCST, Wisconsin Card Sorting Test; WISC, Wechsler Intelligence Scale for Children; WNV, Wechsler Nonverbal Scale

**Table 3.**
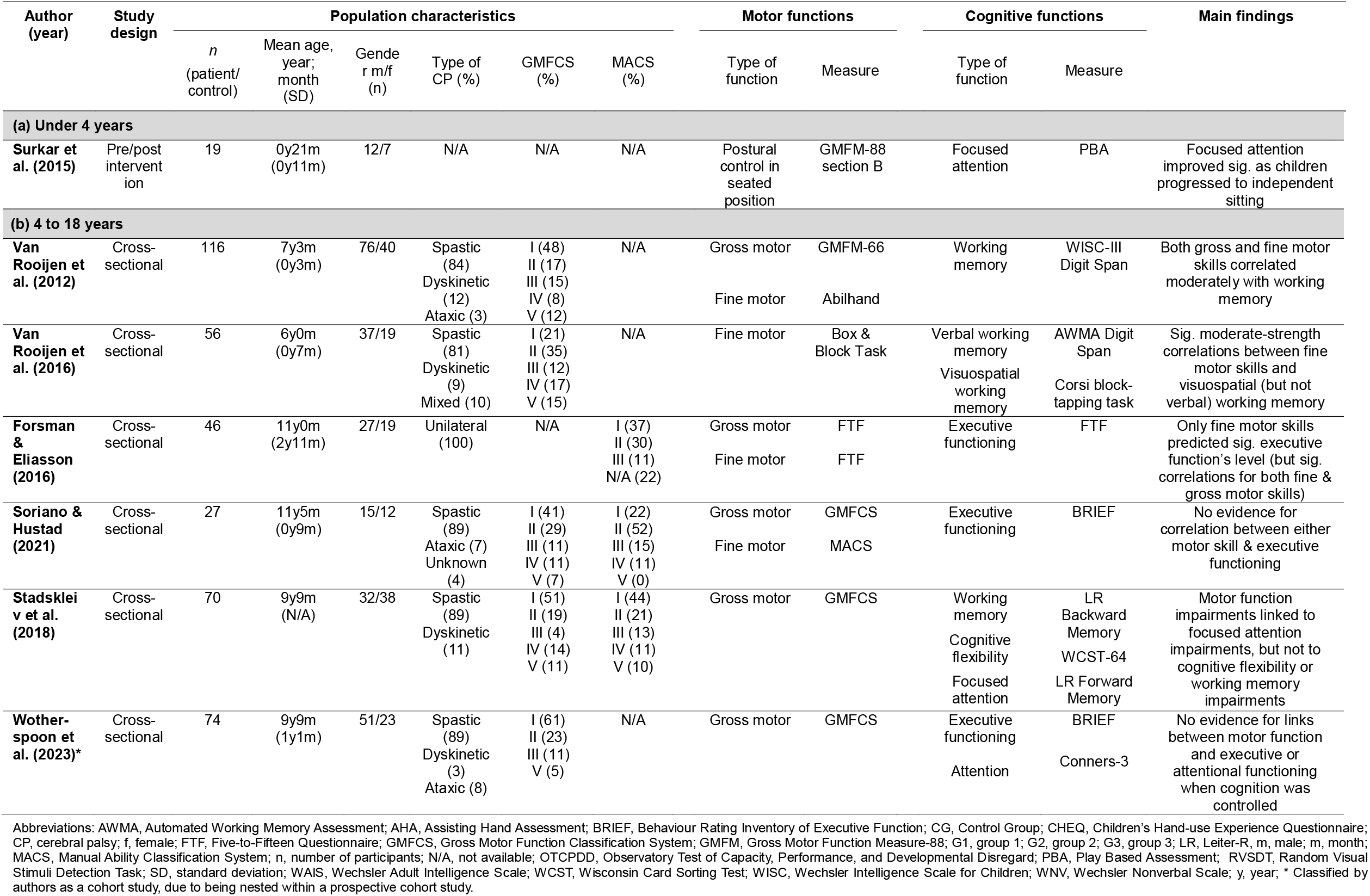
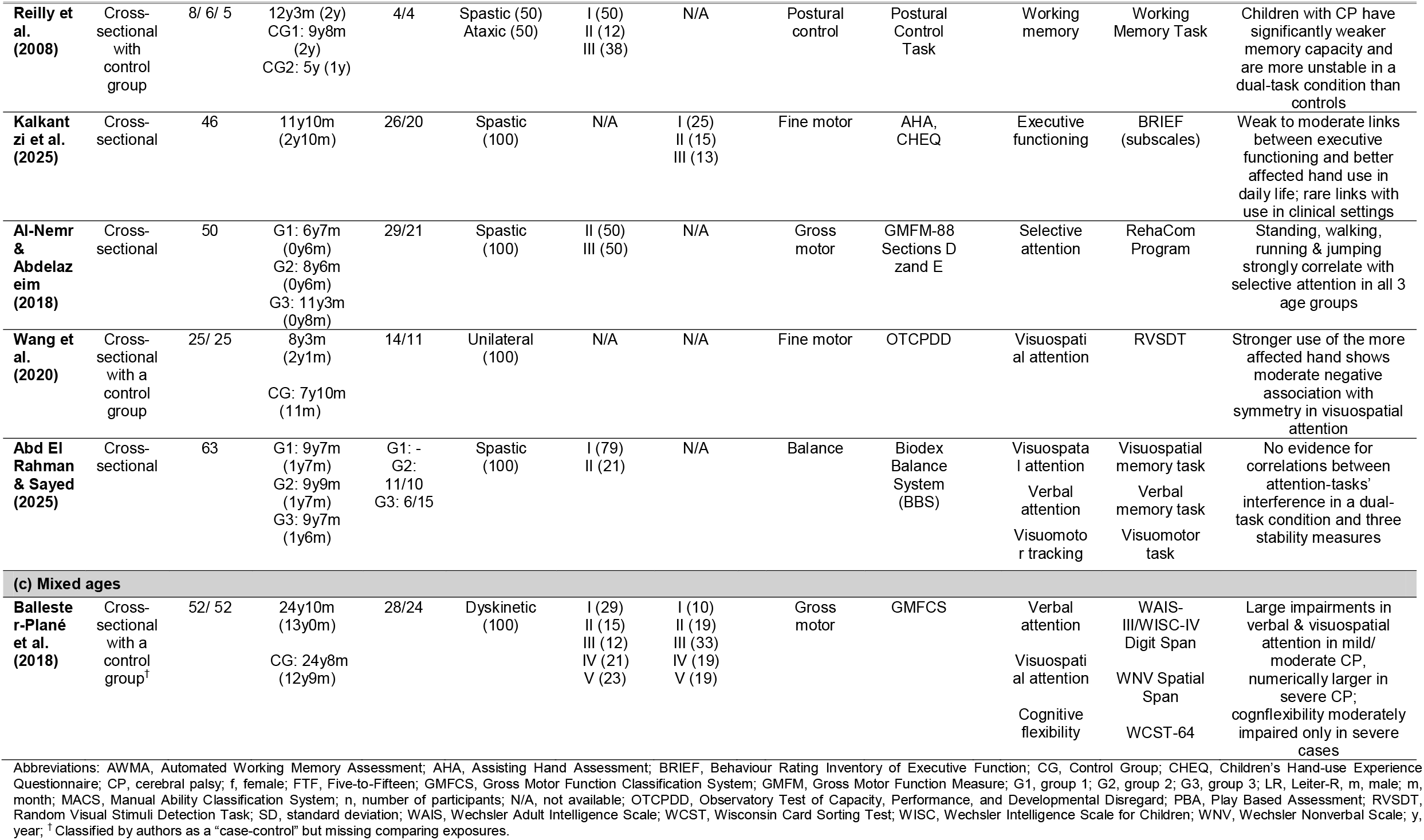
Detailed information from the reviewed studies links between motor and cognitive control functions in CP.

### Population characteristics

Table 3 presents various population characteristics of the 13 reviewed studies. The studies included a total population of 652 participants, aged between 22 months and 62 years. Given the wide age range of participants in the selected studies, the results were divided into three age ranges: under 4 years, 4–18 years old and a mixed-age group, with, respectively, one, eleven and one study in each category. This enabled a more homogeneous and therefore more informative analysis of the relationships between motor and cognitive control functions.

Six studies focused on specific types of CP, other 6 included a mixed sample containing participants with spastic, dyskinetic or ataxic CP, and one study did not include information on the CP subtype. Three studies used both the GMFCS and the MACS to classify the severity of CP impairment, 6 employed only the GMFCS and 2 used only the MACS, while two articles did not report a classification system. Of the 9 articles that applied the GMFCS, 6 included a sample representing all 5 levels, thus allowing the comparison of cross-domain relationships across the full range of motor functioning. Four studies focused on weaker impairments: 1 study was limited to stages I to II, 1 study to I to III, and 2 confined itself to stages II and III. Of the 5 articles using the MACS scale, 3 included a sample representing all 5 levels of the fine motor functioning (from independent manipulation of objects to severe limitation). Two studies focused solely on stages I to III, thus excluding people with more severe fine motor impairments.

### Relationship between motor and cognitive control functions

While we review each study’s approach and findings below, we summarize first the key results. Executive function was tested in 8 articles, of which 3 evaluated its links with both gross and fine motor skills, 4 with gross motor function only, and 2 with fine motor function. Five out of these 8 studies reported significant relationships. Attention was examined in 6 studies, all of which assessed its relationships with gross motor skills, and 4 of whom identified significant correlations (**Error! Reference source not found.3**, *Main findings*). For assessing relationship, correlations (Pearson’s or Spearman’s coefficients) were used in most of the studies, with some using regressions and ANOVAs. Accompanying effects size estimates were not available for 2 studies on attention and 1 on executive functions.

#### Children younger than 4 years

Surkar et al. (2015) examined whether in children with CP under 4 years of age focused attention, measured with a standardized video tool, increased in parallel with gains in postural control measured as ability to sit independently, using the GMFM’s Section B (see Table 2). Following the various interventions aimed at improving seated postural control that all children received during the 8–12 week duration of the study, mean longest periods of focused attention increased as the children moved from supported to independent sitting (*p* < 0.02).

#### Children aged 4 to 18

##### Executive functioning

Five out of the eight studies in this age group showed significant associations. Links with both gross and fine motor skills and executive functions were tested in two studies of predictors of early and more complex arithmetic skills (Van Rooijen et al., 2012, 2016). Fine motor skills showed moderate-size correlations with both visuospatial working memory and updating (*r* = 0.36 and *r* = 0.35, both *p’*s < 0.01) in van Rooijen et al. (2016). Fine motor skills showed also a moderate-size correlation with verbal working memory when the latter was measured with WISC-III (*r* = 0.41, *p* < 0.01; van Rooijen et al. 2012), but not with AWMA (van Rooijen et al. 2016). In van Rooijen et al. (2012) verbal working memory was also significantly and moderately associated with gross motor skills (*r* = 0.40, *p* < 0.01). However, the overall structural equation model, which included fine motor skills, had a better fit without the gross motor skills. Similarly, in Forsman and Eliasson (2016), executive functions were strongly correlated with fine motor skills (*r* = 0.70, *p* < 0.01) and moderately with gross motor skills (*r* = 0.43, *p* < 0.01). However, a linear regression analysis revealed that only fine motor skills (*B* = 0.615, *β* = 0.674, *p* < 0.0001), but not gross motor skills (*B* = 0.088, *β* = 0.096, *p* = 0.42) predicted the level of executive functioning. Again, when fine motor skills were controlled, gross motor skills no longer predicted the level of executive functions.

Two studies here tested one of the two motor skills in isolation. Reilly et al. (2008) compared how children with CP and healthy controls control their posture in a dual task involving a working memory task. Twelve year-olds with CP had lower working memory capacity than age-matched controls. Unlike the age-matched controls, but alike younger, 5-year-old, controls, children with CP experienced dual-task interference in postural control, in multiple stance positions (*p*’s < 0.03). Correlational analyses across all children showed that higher working memory capacity was linked to smaller dual-task postural interference (*r*’s ≥ -0.66, *p*’s < 0.05). Finally, Kalkantzi et al. (2025) showed that in children with unilateral CP general executive functioning is significantly correlated with spontaneous bimanual function when the latter is tested with a parent-reported questionnaire but not with a standardized clinical observation. Almost all of 11 subdimensions of BRIEF questionnaire were negatively correlated, weakly to moderately, with the 3 subdimensions of the bimanual function questionnaire (*r*’s = -0.5 to -0.29; all adjusted *p*’s ≤ 0.05), but only one, weakly, with the clinical observation tool (*r* = -0.35, *p* < 0.05; all n.s. *r*’s < -0.25).

Three of the eight studies in this age group did not find evidence for motor–executive skill associations. Soriano and Hustad (2021) found no evidence for links between general executive functioning, measured with the BRIEF questionnaire, and the fine and gross motor functioning, measured with GMFCS and MACS, respectively (*r* = 0.001, and *r* = 0.209, both *p*’s > 0.05). Also Wotherspoon et al. (2023) failed to show evidence for links between the BRIEF scores and GMFCS levels, when children’s overall cognition was controlled (*p* = 0.81; see below for results for attention). Stadskleiv et al. (2018) in turn failed to find evidence for links between GMFCS and working memory or cognitive flexibility when the latter were measured with computerised tasks (*F*’s < 1.2, *p*’s > 0.3; see below for results for attention).

##### Attention

Three out of five studies in children aged 4 to 18 showed significant associations. Al-Nemr and Abdelazeim (2018) revealed that 6–8-, 8–10- and 10–12-year-olds with CP show significant, strong positive links between task-based selective attention functions and their skills in standing (*r* = 0.87, *r* = 0.73, *r* = 0.71, respectively, all *p’*s < 0.01), and walking, running and jumping (*r* = 0.79, *r* = 0.77, *r* = 0.64, all *p*’s < 0.01). Wang et al. (2020) showed that, children with hemiplegic CP show certain asymmetries in a visual-field spatial attention task compared to age-matched controls. These effects were found mostly in responses misses (overall and peripheral field, *p*’s < 0.02, but not in central field, *p* = 0.99), but not in response times (*p*’s > 0.26). Underuse of the affected hand in these children was negatively and moderately linked to stronger attention asymmetries (again, in response misses, *r* = 0.5, *p* < 0.05, but not response times, *r* = -0.31, *p* n.d.). Finally, the discussed above Stadskleiv et al. (2018) showed that focused attention, measured with standardised tests, in contrast to executive functions, varied significantly with the GMCFS levels (*F* = 6.7, *p* = 0.002).

Two studies in this age group failed to find evidence for motor–attention skill links. Abd El and Sayed (2025) failed to show evidence for correlations between stability and attention measured with three standardized tasks in a dual-task paradigm, *r*’s < 0.1, *p*’s > 0.35. Notably, the above described Wotherspoon et al. (2023) showed that attention ceased to be significantly associated with GMFCS levels when the overall cognitive functioning was controlled (*F* = 1.5, *p* = 0.089).

#### Individuals with CP of mixed ages

In a wide sample of individuals with dyskinetic CP aged 7 to 60 years, Ballester-Plané et al. (2018) showed that GMFCS level is linked to impairments in visuospatial and verbal attention, as well as cognitive flexibility. Pairwise comparisons showed that stronger GMFCS-based motor impairment was associated with stronger and wider impairments in the cognitive functions. Visual and verbal attention were both impaired compared to age- and gender-matched controls already in mild cases. The large impairments were de facto comparable in size across mild and moderate cases (GMFCS I and II/III; r’s = 0.51 to 0.56; all corrected p’s < α = 0.004, beside verbal attention in III/IV, p = 0.005), while being numerically even larger in severe cases, albeit this difference was not statistically tested (GMFCS IV/V; r’s = 0.78 and 0.6, respectively). Notably, only severe cases were also impaired in cognitive flexibility, with this effect being moderate in size (r = 0.47, p = 0.003).

## Discussion

To the best of our knowledge, this is the first review to systematically synthesise the empirical evidence for relationships between motor skills and executive and attentional skills in individuals with CP. A total of 13 studies were included in the review, representing high heterogeneity, in participant characteristics, from their age, CP type, levels of GMFCS and MACS, to the outcome measures and statistical tests used to assess relationships. Nevertheless, the review demonstrates reliable, significant relationships between the two domains in individuals with CP, at both domain-general and process-specific levels.

Executive functions were tested more frequently than attention, but only in the children (4 to 18 years) and mixed-age groups. The strongest evidence was found for a specific function - working memory – which was associated with both fine and gross motor skills (in both cases, 2 out of 3 studies reported significant effects). Similarly, strong evidence was found for overall executive functioning and its association with fine motor skills, whereas only weak evidence was found for associations with gross motor skills (2/3 vs. 1/3). Cognitive flexibility, tested only in association with gross motor skills, also showed mixed evidence (1/2). These associations were mostly all moderate to large size. For attention, significant moderate-to-strong associations were observed across all tested CP age groups. Most evidence was again found for a particular function, here, focused attention, and its associations with gross motor skills (2/2). Gross motor skills showed mixed evidence for links with both verbal and visuospatial attention (1/2 vs. 1/2), single-study evidence of an association with selective attention (1/1), and no evidence of associations with visuomotor attention (0/1) or, notably, general attentional functioning (0/1). Fine motor skills were tested only in one study, showing evidence for links with visuospatial attention, but for only one of the two task measures. Thus, working memory could be a key cognitive control skill in the links with the motor impairments in CP, as it shows reliable strong to moderate associations with both fine and gross motor skills, in both middle and late childhood (Reilly et al. 2008, van Roijen et al. 2012, 2016). Notably, similar strong evidence was found for focused attention, limited to links with gross motor functions, in infancy and late childhood (Stadskleiv et al. 2019; Surkar et al. 2015), and general executive functioning, limited to, in turn, fine motor skills (Forsman & Eliasson, 2016; Kalkantzi et al. 2025).

### Converging evidence for motor–cognitive control associations across populations

Our findings are consistent with the inter-domain links reported in both healthy and other clinical paediatric populations. In typically developing children, the evidence for associations between motor skills and executive functions, at a general level and for specific pairings, is mixed. One large systematic review in 3–12-year-olds showed that balance and manual dexterity were most reliably, weakly, linked to all three basic executive functions (Gandotra et al. 2022). Locomotion was associated only with working memory, and there was no evidence for links involving object control. However, significant weak associations involving executive functions and *all* tested motor competences (general, locomotion, object control, and postural stability) were reported in another large systematic review with a meta-analysis in children and adolescents (Bao et al. 2024). Evidence for these links converged from cross-sectional, longitudinal and experimental studies. In contrast, another systematic review, which directly compared the level of evidence for the inter-domain links in children and adolescents, showed absent or insufficient evidence for many cognitive control–motor associations (van der Fels et al., 2015). Therein, gross motor skills showed strong evidence for *no* links with general executive functioning, insufficient evidence for working memory, and no evidence for associations with general attentional functioning. Fine motor skills showed scant evidence for links with general executive functioning, and no evidence for links with working memory or attention. Although direct comparisons with our findings are difficult due to differences in the associations examined, their strength, and the specific motor–cognitive pairings assessed, our results in children and adolescents with CP similarly indicated associations between general executive functioning and both gross and fine motor skills, as well as working memory. Evidence for associations with cognitive flexibility was mixed, and none of the included studies assessed inhibition. Consistent with van der Fels et al. (2015), we failed to find evidence for links between motor and general attentional functioning. However, in contrast to them, we examined specific attentional functions and identified significant links with gross motor skills, particularly for focused attention, whereas evidence for other attentional functions was more limited. Another notable difference from the findings in the general population was that the motor-cognitive associations observed in CP were predominantly of moderate-to-large magnitude.

Our results broadly parallel the results in developmental coordination disorder (DCD). As in our review in CP, Fogel et al. (2023) reported that in DCD, both fine and gross motor are overall significantly associated, mostly moderately to strongly, with general executive functioning and working memory (links with attention were not evaluated). Complementing this evidence on associations, longitudinal research shows that children at risk of DCD exhibit motor and behavioural difficulties from age three that tend to persist through age five (De Roubaix et al., 2025). Similarly, our results in CP show relatively weaker links of motor functions with attention than with executive functions. These parallels are notable, as both CP and DCD show relatively high co-morbidity with ADHD, 20-30% in CP, and ∼50% for DCD (e.g., Casseus et al. 2024; Gillberg et al. 2004, respectively). In fact, the motor problems in ADHD are proposed to be unidirectionally driven by the core symptoms of ADHD, like executive dysfunction, or caused by under-diagnosed DCD comorbidity (e.g., Goulardins et al. 2017; Damatac et al. 2022; Pant et al. 2022). To what extent the findings of our review are due to potential ADHD comorbidity is largely unclear, warranting caution with the interpretation of results. Out of the 13 included studies, only two recorded ADHD comorbidity (Forsman & Eliasson, 2016; Kalkantzi et al. 2025) and only Kalkantzi et al. (2025) performed sensitivity analyses excluding the ADHD participants. Therein, the weak to moderate links between bimanual function and multiple executive functions that they reported all remained significant, except inhibition. Similarly, attentional impairments in DCD cannot be fully accounted for by co-occurring ADHD, and conversely, motor impairments in ADHD are not fully attributable to DCD comorbidity (e.g., Lee et al. 2021; Lachambre et al. 2021). Thus, although based on a single study and requiring further confirmation, evidence suggests that, in CP and other neuromotor disorders arising from early brain lesions, co-occurring motor and cognitive control impairments may reflect mechanisms that extend beyond ADHD comorbidity.

### Interpretations within a broader context

Our review clarifies the presence and strength of specific inter-domain links from early childhood to adult age. These results are in line with the dominating models of multi-domain neurocognitive development. The dynamic systems theory (Thelen & Smith, 1994; Smith & Thelen, 2003) construes developmental abilities as emerging from reciprocal, non-linear interactions between one’s movements, object interactions and perceptually guided action. Any atypicality in children’s motor capacities would alter how they explore and act on their environment and so modify the opportunities through which they develop attentional and executive functions. Relatedly, the neuronal group selection theory (Hadders-Algra, 2000, 2010) proposes that early brain injury does not trigger simply a single linear pathway from motor impairment to cognitive impairment. Rather, it impairs the repertoire of neural and behavioural solutions available to the child and the process of developing increasingly specialised strategies to cope with demands present in the environment. Within these frameworks, the reduced number and frequency of spontaneous movements observed in CP likely restrict opportunities to explore the environment, learn the sensorimotor consequences of action, and develop motor planning and postural control (Prechtl et al., 1997; Babik et al., 2023). The latter impairments then might restrain or delay the reaching and object knowledge acquisition, delaying or impairing the development of attentional and executive functions. In turn, it would reduce the child’s developing capacities for skilled interactions with a growing number of objects, and related actions.

Our results are in line with both models. First, the presence of reliable motor–cognitive control relationships in CP from early childhood to adult age is consistent with these interlinks representing dynamic, reciprocal interactions throughout development, rather than a uniform, unidirectional consequence of an early motor-to-cognitive cascade. This notion is supported by, for example, strong attentional impairments even in mild CP cases, and more severe cases possessing additional impairments in cognitive flexibility (Ballester-Plané et al., 2018). Our results also show that the motor–cognitive relationships in CP are not global but instead vary systematically according to the specific motor and cognitive processes. In this, they enrich both models, which do not propose explicitly that the links may be specific to distinct functions across the two domains. Specifically, we show relatively wide motor associations with working memory, more selective ones for focused attention, and relatively mixed results for general attentional functioning and cognitive flexibility. This pattern also suggests that the developmental coupling might be stronger for functions supporting the maintenance, selection and organisation of action-relevant information. However, due to the largely observational nature of all our reviewed studies, we cannot yet distinguish between the shared, injury-origin versus genuinely reciprocal nature of the underlying mechanisms.

Taken together, a clearer picture of CP as a developmental condition emerges. The weaker inter-domain links observed in CP than in DCD suggest that reciprocal motor–cognitive control interactions may contribute less to the clinical phenotype of CP than to that of the DCD. The early brain injuries of the developing brain likely affect both motor and cognitive control systems as a common neural cause. Nevertheless, motor-to-cognitive influences should remain important, as constrained movements would reduce exploration, independent action and interaction with the environment, as well as learning opportunities and adaptation. Attention-to-motor influences would likewise be important, for example in shaping higher-level motor skills and everyday performance. However, in CP, both motor and attentional impairments are likely to be secondary consequences of the underlying brain lesion. As a result, cross-domain coupling may be weaker than in DCD or ADHD, where motor and cognitive difficulties may reflect shared developmental disruptions in sensorimotor processes and environmental exploration (e.g., Shimko & James, 2025). Evidence consistent with motor–attention developmental cascades also comes from infant siblings at elevated likelihood of autism, in whom motor skills at 10 months predicted later language abilities partly through joint attention at 14 months (Bruyneel et al., 2019). An important outstanding question for theories of cross-domain development is whether motor and cognitive impairments remain similarly coupled when the primary disruption affects the sensory system. A related question is whether the strength or nature of these associations differs between severe sensory impairments, such as early blindness, where vision is largely or entirely unavailable, and milder impairments, such as amblyopia, where atypical early visual experience alters neurodevelopment (e.g., Murray et al. 2015; Simon-Martinez et al. 2023).

### Future research

Development proceeds through long-term dynamic interactions across the system implicating the brain, the body and the behaviour (Byrge et al. 2014). To gain a clearer understanding of the developmental trajectories and the interactions across the two domains in CP, we need more longitudinal studies that track motor and cognitive control development from infancy throughout to adulthood, using multiple levels of evidence. Until now, virtually all studies (beside one, using an intervention) evaluating these inter-links employed cross-sectional designs. Longitudinal studies would help verify if the accounts of Thelen and Smith (1994), and Hadders-Algra (2000, 2010) apply also to CP. Specifically, they would allow to test if the inter-links between motor and cognitive impairments emerge from a cascade of atypical dynamic, reciprocal cross-domain interactions throughout childhood (Karmiloff-Smith, 1997), in which cascade the brain injury is only or mostly the trigger. In this case, the injury-driven interruptions in the action control would alter how these children interact, refine cognitive skills towards and adapt to their environments (moto-to-cognitive influences), and, later, the delayed or impaired cognitive (control) skills would impoverish subsequent capacities to develop more complex motor skills, like dexterity (motor-to-attention influences). This could manifest as a gradual decline in the ability of brain injury characteristics, such as lesion extent or location, to predict later outcomes, whereas the predictive power of previously acquired motor and cognitive skills for subsequent skill acquisition increases over development. Alternatively, since the degree of the motor dysfunction is strongly related to the early brain injury in CP, the motor–cognitive interlinks in CP may instead be predominantly the parallel consequences of the early brain injury. In this case, the brain injury characteristics, like the location and/or extent, would remain the main and stable predictor of both motor and cognitive functioning throughout development. Differences in lesion location and severity could in principle explain the substantial variability we observed in the presence and strength of associations between the two domains. In other words, the characteristics of the brain lesions also need to be recorded by the future longitudinal studies. Observing how the competences across both domains emerge and stabilise during key developmental transitions could also provide converging evidence regarding whether motor and cognitive impairments represent largely independent consequences of the early brain injury, mediated by early motor atypicalities, or instead reflect interconnected developmental adaptations (Kretch et al., 2024).

To strengthen their conclusions, future research should use standardised measures within each domain, moving towards universal assessment protocols that can accommodate the wide range of motor and cognitive abilities observed in individuals with CP. Such protocols would facilitate the systematic evaluation of cross-domain associations across specific motor phenotypes (e.g. spastic, dystonic), and across the full spectrum of upper and lower extremity function. With this, future studies would be able to establish the specificity and selectivity of the inter-domain links as a function of the type and level of motor (dys)function. Healthy controls, matched on relevant dimensions, should also be tested, to understand normative developmental patterns from those that are specific to CP. To better understand shared and distinct co-developmental mechanisms across neurodevelopmental conditions, future studies should systematically assess and account for co-occurring ADHD and autism, given their partly overlapping but non-identical profiles of executive-function difficulties (e.g., Geurts et al., 2004). More generally, future research should use rich, multi-modal and longitudinal approaches, to assess changes in motor function, cognitive skills, and neural responses across time. Such designs are needed to shed light on (co-)developmental trajectories, identify crucial transitional periods, and characterize the stability and plasticity of motor, cognitive, and sensory systems over time (e.g., Maitre et al. 2017; Matusz et al. 2018). Crucially, these processes should be studied where development and learning occur, i.e., in naturalistic contexts such as home, playground or classroom (e.g., Matusz et al. 2019; Marschik et al. 2023). Consistently, classroom observations show that differences in on-task behaviour between children with ADHD and their peers vary with task demands and instructional setting, e.g., across individual, small group and whole class work (Imeraj et al., 2013). In such environments, evidence from standardized tests can be complemented by more naturalistic, dynamic and complex measures, provided by mobile EEG/fNIRS or head-mounted eye-tracking, momentary self-reports or movement sensors, and supported by machine learning methods for processing and analysis. Such objective measures are important to offer a more accurate picture on child’s functioning, which caregiver reports are known to underestimate (Ramstad et al. 2015; Birnie et al. 2020).

### Implications for practice

Our review shows that motor and cognitive functions are indeed connected in individuals with CP. These findings highlight the importance, and its increasing recognition by the field, of intervening into not only motor, but also sensory and cognitive functions (especially cognitive control), due to the importance of latter for development and daily functioning (Stadskleiv et al. 2020; Fluss & Lidzba, 2020). Our review also extends findings of reviews of interventional studies, by providing, observational in nature, evidence that in CP the links between motor and cognitive control encompass also executive functions for which experimental evidence is mixed (Blasco et al. 2023; Carracedo-Martín et al., 2025). Our results suggest that more comprehensive clinical profiling spanning both domains might be necessary to identify the needs of individuals with CP, as the severity of motor difficulties may also signal the presence of attentional or executive impairments (e.g., Ballester-Plané et al., 2018). Such more comprehensive phenotyping should support designing better tailored interventions, targeting the broader functional profile of CP (e.g., Marschik et al. 2017). For children with more severe motor impairments, who are often excluded from cognitive evaluations because conventional testing methods can be difficult to administer (Soriano & Hustad, 2021; Stadskleiv et al., 2018), emerging technologies, like wearables devices or mobile neuroimaging may offer new opportunities to assess cognitive capacities. Nevertheless, further research is needed to determine whether there are individuals with CP in whom the severity of motor and cognitive control impairments is unrelated and, if so, how common such profiles are. These individuals may differ in characteristics of their early-life brain injury, such as lesion location or extent. Such knowledge would inform clinical decision-making by clarifying the extent to which interventions targeting one domain may also benefit the other.

The importance of early intervention in CP is well-established (e.g. Morgan et al. 2021; Spittle & Morgan 2025), due to the cascading and large effects it has on subsequent, long-term development. In line with the dynamic systems theory, even small changes in the child’s environment during sensitive developmental periods should have extensive effects across multiple developmental domains (Thelen & Smith, 1994). At the same time, recent literature reviews demonstrate that physical or mixed-activity interventions, such as aerobic or strength training, yoga, meditation- or dance-based approaches, in populations largely consisting of children and adults, have reliable benefits for attention, and more variable effects for executive functions (Blasco et al. 2023; Carracedo-Martín et al., 2025). Physical activity interventions improve executive and attentional functions in typical and ADHD populations (Benzing et al., 2018; de Greeff et al., 2018; Zhu et al., 2023). Most of our reviewed studies also focused on these two age groups, and, albeit the observational nature of these results cannot offer causal evidence, our review provides converging, indirect evidence for such relationships. This is important, as physical activity or exergames are engaging, cost-effective and easy to integrate in everyday life, and as such could have important wide-range benefits also for individuals with CP from childhood onwards.

## Conclusion

This scoping review synthesises evidence for associations between motor function, and executive and attentional abilities in individuals with CP. Across 13 heterogeneous studies, the findings suggest that these relationships vary according to the specific motor and cognitive control functions assessed. The most consistent evidence concerned associations of working memory with both fine and gross motor skills, focused attention with gross motor skills, and general executive functioning with fine motor skills, although each was supported by only a small number of studies. These patterns are compatible with developmental frameworks emphasising dynamic interactions between motor and cognitive functions. However, the predominantly cross-sectional nature of the reviewed evidence prevents establishing the direction or causal nature of these relationships, or distinguishing between shared consequences of early brain injury and reciprocal developmental influences. Longitudinal studies incorporating lesion characteristics, relevant comorbidities, and accessible, standardised assessments are needed to clarify these mechanisms and their developmental trajectories. Intervention trials should test whether improvements in one domain produce benefits in the other. Overall, these findings support the comprehensive assessment of both motor and cognitive control functions to inform individualised, interdisciplinary care. They also provide a rationale for investigating interventions addressing both domains, while highlighting that cross-domain benefits remain to be established.

## Data Availability

This is a systematic review paper and as such the paper did not produce original data that could be shared.

## Declarations

For this work, we affirm that there is no known conflict of interest. None of the co-authors had any financial or non-financial interests linked to completing this work.

## Acknowledgements

This study is based on the work prepared by MM and KR for a Bachelor’s degree in Physiotherapy at the Physiotherapy Section of School of Health Sciences, HES-SO Valais, under the direction of PJM, and developed further as part of a collaboration conducted with JS, DZ and PBM. PBM is funded by the Federal Ministry of Research, Technology and Space (Bundesministerium für Forschung, Technologie und Raumfahrt; BMFTR) as part of the German Center for Child and Adolescent Health (DZKJ) under the funding code 01GL2402A.

## Supplementary material

## Appendix 1

**Table 2.**
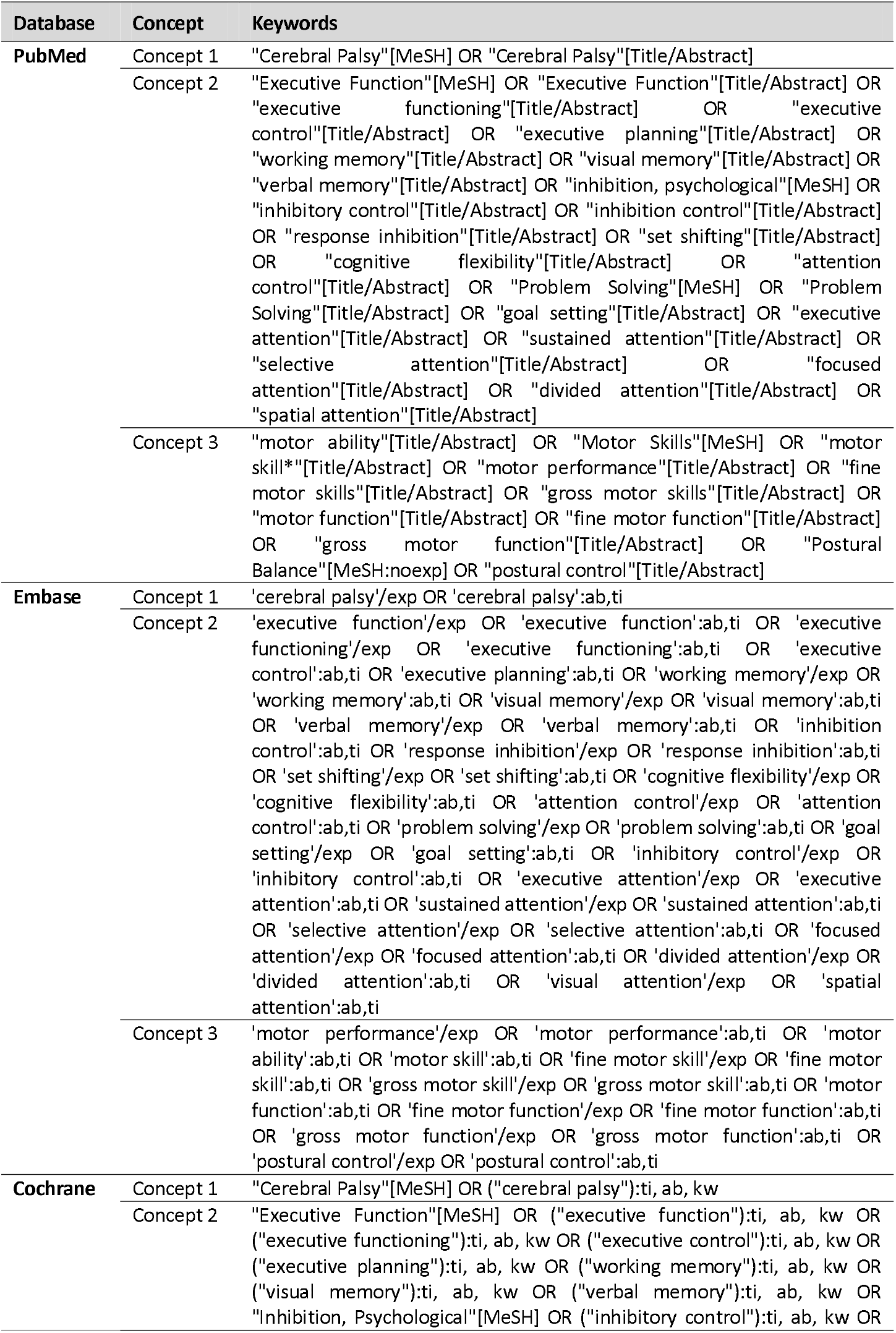
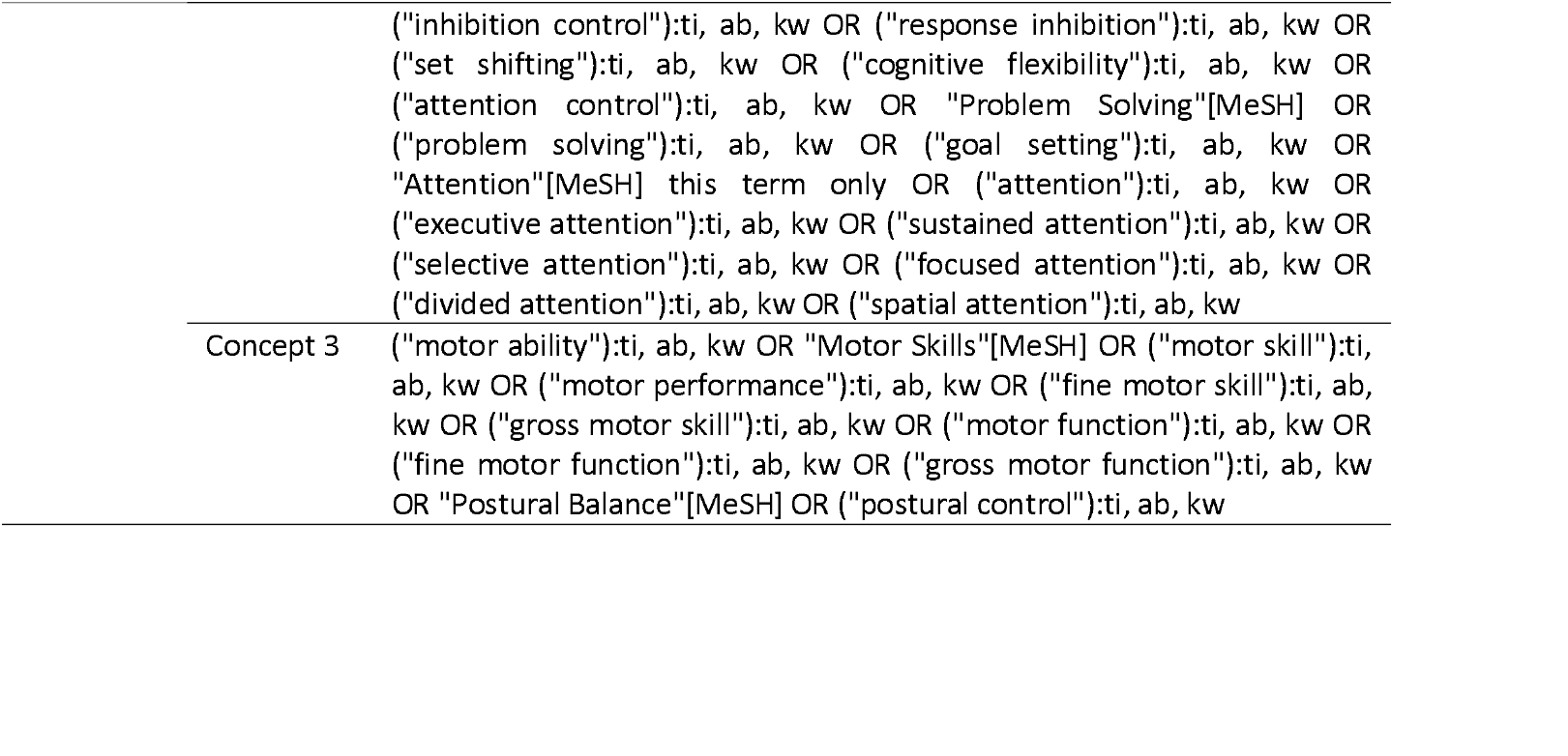
Research equation.

| Database | Concept | Keywords |
| --- | --- | --- |
| PubMed | Concept 1 | "Cerebral Palsy"[MeSH] OR "Cerebral Palsy"[Title/Abstract] |
|  | Concept 2 | "Executive Function"[MeSH] OR "Executive Function"[Title/Abstract] OR "executive functioning"[Title/Abstract] OR "executive control"[Title/Abstract] OR "executive planning"[Title/Abstract] OR "working memory"[Title/Abstract] OR "visual memory"[Title/Abstract] OR "verbal memory"[Title/Abstract] OR "inhibition, psychological"[MeSH] OR "inhibitory control"[Title/Abstract] OR "inhibition control"[Title/Abstract] OR "response inhibition"[Title/Abstract] OR "set shifting"[Title/Abstract] OR "cognitive flexibility"[Title/Abstract] OR "attention control"[Title/Abstract] OR "Problem Solving"[MeSH] OR "Problem Solving"[Title/Abstract] OR "goal setting"[Title/Abstract] OR "executive attention"[Title/Abstract] OR "sustained attention"[Title/Abstract] OR "selective attention"[Title/Abstract] OR "focused attention"[Title/Abstract] OR "divided attention"[Title/Abstract] OR "spatial attention"[Title/Abstract] |
|  | Concept 3 | "motor ability"[Title/Abstract] OR "Motor Skills"[MeSH] OR "motor skill*"[Title/Abstract] OR "motor performance"[Title/Abstract] OR "fine motor skills"[Title/Abstract] OR "gross motor skills"[Title/Abstract] OR "motor function"[Title/Abstract] OR "fine motor function"[Title/Abstract] OR "gross motor function"[Title/Abstract] OR "Postural Balance"[MeSH:noexp] OR "postural control"[Title/Abstract] |
| Embase | Concept 1 | 'cerebral palsy'/exp OR 'cerebral palsy':ab,ti |
|  | Concept 2 | 'executive function'/exp OR 'executive function':ab,ti OR 'executive functioning'/exp OR 'executive functioning':ab,ti OR 'executive control':ab,ti OR 'executive planning':ab,ti OR 'working memory'/exp OR 'working memory':ab,ti OR 'visual memory'/exp OR 'visual memory':ab,ti OR 'verbal memory'/exp OR 'verbal memory':ab,ti OR 'inhibition control':ab,ti OR 'response inhibition'/exp OR 'response inhibition':ab,ti OR 'set shifting'/exp OR 'set shifting':ab,ti OR 'cognitive flexibility'/exp OR 'cognitive flexibility':ab,ti OR 'attention control'/exp OR 'attention control':ab,ti OR 'problem solving'/exp OR 'problem solving':ab,ti OR 'goal setting'/exp OR 'goal setting':ab,ti OR 'inhibitory control'/exp OR 'inhibitory control':ab,ti OR 'executive attention'/exp OR 'executive attention':ab,ti OR 'sustained attention'/exp OR 'sustained attention':ab,ti OR 'selective attention'/exp OR 'selective attention':ab,ti OR 'focused attention'/exp OR 'focused attention':ab,ti OR 'divided attention'/exp OR 'divided attention':ab,ti OR 'visual attention'/exp OR 'spatial attention':ab,ti |
|  | Concept 3 | 'motor performance'/exp OR 'motor performance':ab,ti OR 'motor ability':ab,ti OR 'motor skill':ab,ti OR 'fine motor skill'/exp OR 'fine motor skill':ab,ti OR 'gross motor skill'/exp OR 'gross motor skill':ab,ti OR 'motor function':ab,ti OR 'fine motor function'/exp OR 'fine motor function':ab,ti OR 'gross motor function'/exp OR 'gross motor function':ab,ti OR 'postural control'/exp OR 'postural control':ab,ti |
| Cochrane | Concept 1 | "Cerebral Palsy"[MeSH] OR ("cerebral palsy"):ti, ab, kw |
|  | Concept 2 | "Executive Function"[MeSH] OR ("executive function"):ti, ab, kw OR ("executive functioning"):ti, ab, kw OR ("executive control"):ti, ab, kw OR ("executive planning"):ti, ab, kw OR ("working memory"):ti, ab, kw OR ("visual memory"):ti, ab, kw OR ("verbal memory"):ti, ab, kw OR "Inhibition, Psychological"[MeSH] OR ("inhibitory control"):ti, ab, kw OR |

|  |  |
| --- | --- |
|  | ("inhibition control"):ti, ab, kw OR ("response inhibition"):ti, ab, kw OR<br>("set shifting"):ti, ab, kw OR ("cognitive flexibility"):ti, ab, kw OR<br>("attention control"):ti, ab, kw OR "Problem Solving"[MeSH] OR<br>("problem solving"):ti, ab, kw OR ("goal setting"):ti, ab, kw OR<br>"Attention"[MeSH] this term only OR ("attention"):ti, ab, kw OR<br>("executive attention"):ti, ab, kw OR ("sustained attention"):ti, ab, kw OR<br>("selective attention"):ti, ab, kw OR ("focused attention"):ti, ab, kw OR<br>("divided attention"):ti, ab, kw OR ("spatial attention"):ti, ab, kw |
| Concept 3 | ("motor ability"):ti, ab, kw OR "Motor Skills"[MeSH] OR ("motor skill"):ti,<br>ab, kw OR ("motor performance"):ti, ab, kw OR ("fine motor skill"):ti, ab,<br>kw OR ("gross motor skill"):ti, ab, kw OR ("motor function"):ti, ab, kw OR<br>("fine motor function"):ti, ab, kw OR ("gross motor function"):ti, ab, kw<br>OR "Postural Balance"[MeSH] OR ("postural control"):ti, ab, kw |

## Footnotes

1 The Manual Ability Classification System (MACS) classifies across five levels how children (4–18 years) with CP use their hands to handle objects in daily activities.

## Notes

### Competing Interest Statement

The authors have declared no competing interest.

### Summary of Updates

We have integrated two internationally renowned experts in cerebral palsy, Dre Dajie Zhang and Prof. Peter B. Marschik, as co-authors, in order to improve the clinical and research quality of the developed review, and so the offered conclusions.

## References

Al-Nemr, A., & Abdelazeim, F. (2018). Relationship of cognitive functions and gross motor abilities in children with spastic diplegic cerebral palsy. Applied Neuropsychology: Child, 7(3), 268–276. 10.1080/21622965.2017.1312402 Abd El, M. A. E. N. R., & Sayed, A. O. (2025). Effect of Disturbed Attention-Demanding Tasks on Balance in Children with Spastic Cerebral Palsy: Cross-Sectional Study. The Egyptian Journal of Hospital Medicine, 100, 3452–3455.

Arnould, C., Penta, M., Renders, A., & Thonnard, J. L. (2004). ABILHAND-Kids: a measure of manual ability in children with cerebral palsy. Neurology, 63(6), 1045–1052. 10.1212/01.WNL.0000138423.77640.3

Babik, I., Cunha, A. B., & Srinivasan, S. (2023). Biological and environmental factors may affect children’s executive function through motor and sensorimotor development: Preterm birth and cerebral palsy. Infant Behavior and Development, 73, 101881. 10.1016/j.infbeh.2023.101881

Ballester-Plané, J., Laporta-Hoyos, O., Macaya, A., Póo, P., Meléndez-Plumed, M., Toro-Tamargo, E., Gimeno, F., Narberhaus, A., Segarra, D., & Pueyo, R. (2018). Cognitive functioning in dyskinetic cerebral palsy: Its relation to motor function, communication and epilepsy. European Journal of Paediatric Neurology, 22(1), 102–112. 10.1016/j.ejpn.2017.10.006

Bao, R., Wade, L., Leahy, A. A., Owen, K. B., Hillman, C. H., Jaakkola, T., & Lubans, D. R. (2024). Associations between motor competence and executive functions in children and adolescents: A systematic review and meta-analysis. Sports Medicine. 10.1007/s40279-024-02040-1.

Benzing, V., Chang, Y. K., & Schmidt, M. (2018). Acute physical activity enhances executive functions in children with ADHD. Scientific reports, 8(1), 12382. 10.1038/s41598-018-30067-8.

Bernardi, M., Leonard, H. C., Hill, E. L., Botting, N., & Henry, L. A. (2018). Executive functions in children with developmental coordination disorder: a 2-year follow-up study. Developmental Medicine & Child Neurology, 60(3), 306–313. 10.1111/dmcn.13640

Best, J. R., & Miller, P. H. (2010). A developmental perspective on executive function. Child Development, 81(6), 1641–1660. 10.1111/j.1467-8624.2010.01499.x

Birnie, K.A., Richardson, P.A., Rajagopalan, A.V., Bhandari, R.P. (2020). Factors related to agreement between child and caregiver report of child functioning with chronic pain. Clin. J. Pain, 36, 203–212. 10.1097/AJP.0000000000000794.

Blasco, M., García-Galant, M., Berenguer-González, A., Caldú, X., Arqué, M., Laporta-Hoyos, O., & Pueyo, R. (2023). Interventions with an impact on cognitive functions in cerebral palsy: a systematic review. Neuropsychology review, 33(2), 551–577.

Bottcher, L. (2010). Children with spastic cerebral palsy, their cognitive functioning, and social participation: a review. Child neuropsychology, 16(3), 209–228.

Bottcher, L., Flachs, E. M., & Uldall, P. (2010). Attentional and executive impairments in children with spastic cerebral palsy. Developmental Medicine & Child Neurology, 52(2), e42–e47.

Bruyneel, E., Demurie, E., Warreyn, P., & Roeyers, H. (2019). Study of joint attention as a mediator between motor skills and receptive/expressive language in infant siblings at risk for autism. Infant Behavior and Development, 57, 101377. 10.1016/j.infbeh.2019.101377

Byrge, L., Sporns, O., & Smith, L. B. (2014). Developmental process emerges from extended brain–body– behavior networks. Trends in Cognitive Sciences, 18, 395–403.

Cabezas, M., & Carriedo, N. (2020). Inhibitory control and temporal perception in cerebral palsy. Child Neuropsychology, 26(3), 362–387. 10.1080/09297049.2019.1656712

Calignano, G., Lettere, G., Leo, I., Maritan, F., Mattera, L., Granata, P., Lucangeli, D., & Valenza, E. (2023). Interconnected Pathways: Postural Stability and Vocabulary Skills in Preschool-Aged Children. Children, 10. 10.3390/children10121891.

Carracedo-Martín, M., Moral-Salicrú, P., Blasco, M., Fernández-Andújar, M., Pueyo, R., & Ballester-Plané, J. (2025). The Role of Cognitive Functioning in the ICF Framework: A Systematic Review of Its Influence on Activities and Participation and Environmental Factors in People with Cerebral Palsy. Journal of Clinical Medicine, 14(18), 6393. 10.3390/jcm14186393.

Casseus, M., Cheng, J., & Reichman, N. E. (2024). Clinical and functional characteristics of children and young adults with cerebral palsy and co-occurring attention-deficit/hyperactivity disorder. Research in developmental disabilities, 151, 104787. doi: 10.1016/j.ridd.2024.104787.

Crichton, A., Ditchfield, M., Gwini, S., Wallen, M., Thorley, M., Bracken, J., Harvey, A., Elliott, C., Novak, I., & Hoare, B. (2020). Brain magnetic resonance imaging is a predictor of bimanual performance and executive function in children with unilateral cerebral palsy. Developmental Medicine and Child Neurology, 62(5), 615–624. 10.1111/dmcn.14462

Damatac, C. G., Soheili-Nezhad, S., Freches, G. B., Zwiers, M. P., De Bruijn, S., Ikde, S., … & Sprooten, E. (2022). Longitudinal changes of ADHD symptoms in association with white matter microstructure: A tract-specific fixel-based analysis. NeuroImage: Clinical, 35, 103057. 10.1016/j.nicl.2022.103057

De Greeff, J. W., Bosker, R. J., Oosterlaan, J., Visscher, C., & Hartman, E. (2018). Effects of physical activity on executive functions, attention and academic performance in preadolescent children: a meta-analysis. Journal of science and medicine in sport, 21(5), 501–507. 10.1016/j.jsams.2017.09.595

De Roubaix, A., Dewitte, G., Vens, N., Faes, F., Zwicker, J. G., Dan, B., & Van Waelvelde, H. (2025). Motor, physical, and behavioural performance of 3-to 5-year-old children at risk of developmental coordination disorder: A longitudinal observational study. Developmental Medicine & Child Neurology, 67(12), 1601–1615. 10.1111/dmcn.16370.

Diamond, A. (2000). Close interrelation of motor development and cognitive development and of the cerebellum and prefrontal cortex. Child Development, 71(1), 44–56. 10.1111/1467-8624.00117

Dusing, S. C., & Harbourne, R. T. (2010). Variability in Postural Control During Infancy: Implications for Development, Assessment, and Intervention. Physical Therapy, 90(12), 1838–1849. 10.2522/ptj.2010033

Einspieler, C., Prayer, D., & Marschik, P. B. (2021). Fetal movements: the origin of human behaviour. Developmental Medicine & Child Neurology, 63(10), 1142–1148.

Einspieler, C., Bos, A. F., Libertus, M. E., & Marschik, P. B. (2016). The General Movement Assessment Helps Us to Identify Preterm Infants at Risk for Cognitive Dysfunction. Frontiers in Psychology, 7. 10.3389/fpsyg.2016.00406.

Einspieler, C., Marschik, P. B., & Prechtl, H. F. (2008). Human motor behavior: Prenatal origin and early postnatal development. Journal of Psychology, 216(3), 147–153.

Eliasson, A. C., Krumlinde-Sundholm, L., Rösblad, B., Beckung, E., Arner, M., Öhrvall, A. M., & Rosenbaum, P. (2006). The Manual Ability Classification System (MACS) for children with cerebral palsy: scale development and evidence of validity and reliability. Developmental medicine and child neurology, 48(7), 549–554. 10.1017/S0012162206001162

Fluss, J., & Lidzba, K. (2020). Cognitive and academic profiles in children with cerebral palsy: A narrative review. Annals of physical and rehabilitation medicine, 63(5), 447–456. 10.1016/j.rehab.2020.01.005.

Fogel, Y., Stuart, N., Joyce, T., & Barnett, A. L. (2023). Relationships between motor skills and executive functions in developmental coordination disorder (DCD): A systematic review. Scandinavian Journal of Occupational Therapy, 30(3), 344–356. 10.1080/11038128.2021.2019306

Forsman, L., & Eliasson, A.-C. (2016). Strengths and challenges faced by school-aged children with unilateral CP described by the Five To Fifteen parental questionnaire. Developmental Neurorehabilitation, 19(6), 380–388. 10.3109/17518423.2015.1017662

Gabbard, C. (2008). Lifelong Motor Development. Pearson Benjamin Cummings.

Gandotra, A., Kótyuk, S., Sattar, Y., Bizonics, V., Csaba, R., Cserényi, R., & Cserjesi, E. (2021). A meta-analysis of the relationship between motor skills and executive functions in typically-developing children. Journal of Cognition and Development, 23(8), 1–28. 10.1080/15248372.2021.1979554

Geurts, H. M., Verté, S., Oosterlaan, J., Roeyers, H., & Sergeant, J. A. (2004). How specific are executive functioning deficits in attention deficit hyperactivity disorder and autism? Journal of Child Psychology and Psychiatry, 45(4), 836–854. 10.1111/j.1469-7610.2004.00276.x

Gillberg, C., Gillberg, I. C., Rasmussen, P., Kadesjö, B., Söderström, H., Råstam, M., & Niklasson, L. (2004). Co– existing disorders in ADHD–implications for diagnosis and intervention. European child & adolescent psychiatry, 13(Suppl 1), i80–i92.

Gioia, G. A., Isquith, P. K., Retzlaff, P. D., & Espy, K. A. (2002). Confirmatory factor analysis of the Behavior Rating Inventory of Executive Function (BRIEF) in a clinical sample. Child neuropsychology, 8(4), 249–257. doi: 10.1076/chin.8.4.249.13513. https://doi.org/10.1076/chin.8.4.249.13513.

Goulardins, J. B., Marques, J. C., & De Oliveira, J. A. (2017). Attention deficit hyperactivity disorder and motor impairment: A critical review. Perceptual and motor skills, 124(2), 425–440. 10.1177/0031512517690607.

Hadders-Algra, M. (2010). Variation and variability: key words in human motor development. Physical therapy, 90(12), 1823–1837. 10.2522/ptj.20100006.

Hadders-Algra, M. (2000). The neuronal group selection theory: a framework to explain variation in normal motor development. Developmental medicine and child neurology, 42(8), 566–572. 10.1017/S0012162200001067.

Houwen, S., Visser, L., van der Putten, A., & Vlaskamp, C. (2016). The interrelationships between motor, cognitive, and language development in children with and without intellectual and developmental disabilities. Research in developmental disabilities, 53, 19–31. doi: 10.1016/j.ridd.2016.01.012.

Imeraj, L., Antrop, I., Sonuga-Barke, E., Deboutte, D., Deschepper, E., Bal, S., & Roeyers, H. (2013). The impact of instructional context on classroom on-task behavior: A matched comparison of children with ADHD and non-ADHD classmates. Journal of School Psychology, 51(4), 487–498. 10.1016/j.jsp.2013.05.004.

Kadesjö, B., Janols, L. O., Korkman, M., Mickelsson, K., Strand, G., Trillingsgaard, A., & Gillberg, C. (2004). The FTF (Five to Fifteen): the development of a parent questionnaire for the assessment of ADHD and comorbid conditions. European child & adolescent psychiatry, 13(Suppl 3), iii3–iii13.

Kasap, Z., Çobanoğlu Osmanlı, C., Sarı, İ. F., Er, E., Şahin, B., & Kulaklı, F. (2024). Exploring the quantity and quality of symptoms of attention deficit hyperactivity disorder and intelligence in children with cerebral palsy: a case–control study. European journal of pediatrics, 183(11), 4905–4912. 10.1007/s00431-024-05740-y

Karimi, N., Ebrahimi, I., Kahrizi, S., & Torkaman, G. (2008). Evaluation of postural balance using the biodex balance system in subjects with and without low back pain. Pakistan Journal of Medical Sciences, 24(3), 372.

Kalkantzi, A., Kleeren, L., Baeyens, D., Decraene, L., Crotti, M., Klingels, K., & Mailleux, L. (2025). Daily-life executive functions and bimanual performance in children with unilateral cerebral palsy. Developmental Medicine & Child Neurology, 67, 1290–1300.

Karmiloff-Smith, A. (1997). Crucial differences between developmental cognitive neuroscience and adult neuropsychology. Developmental neuropsychology, 13(4), 513–524. 10.1080/87565649709540693.

Klupp, S., Möhring, W., Lemola, S., & Grob, A. (2021). Relations between fine motor skills and intelligence in typically developing children and children with attention deficit hyperactivity disorder. Research in developmental disabilities, 110, 103855. doi: 10.1016/j.ridd.2021.103855.

Kretch, K. S., Koziol, N. A., Marcinowski, E. C., Hsu, L. Y., Harbourne, R. T., Lobo, M. A., … & Dusing, S. C. (2024). Sitting capacity and performance in infants with typical development and infants with motor delay. Physical & occupational therapy in pediatrics, 44(2), 164–179.

Krumlinde-Sundholm, L. & Eliasson, A.C. (2003). Development of the Assisting Hand Assessment: A Rasch-built Measure intended for Children with Unilateral Upper Limb Impairments. Scandinavian Journal of Occupational Therapy, 10.

Kyvelidou, A., Koss, K., Wickstrom, J., Needelman, H., Fisher, W. W., & DeVeney, S. (2021). Postural control may drive the development of other domains in infancy. Clinical biomechanics, 82, 105273. 10.1016/j.clinbiomech.2021.105273

Lachambre, C., Proteau-Lemieux, M., Lepage, J. F., Bussieres, E. L., & Lippe, S. (2021). Attentional and executive functions in children and adolescents with developmental coordination disorder and the influence of comorbid disorders: A systematic review of the literature. PloS one, 16(6), e0252043.

Lee, J., Mayall, L. A., Bates, K. E., Hill, E. L., Leonard, H. C., & Farran, E. K. (2021). The relationship between motor milestone achievement and childhood motor deficits in children with Attention Deficit Hyperactivity Disorder (ADHD) and children with Developmental Coordination Disorder. Research in Developmental Disabilities, 113, 103920

Libertus, K., & Violi, D. A. (2016). Sit to Talk: Relation between Motor Skills and Language Development in Infancy. Frontiers in Psychology, 7. 10.3389/fpsyg.2016.00475

Lodha, S., & Gupta, R. (2022). Mindfulness, attentional networks, and executive functioning: A review of interventions and long-term meditation practice. Journal of Cognitive Enhancement, 6(4), 531–548. 10.1007/s41465-022-00254-7

Maioli, C., Falciati L, Galli J, Micheletti S, Turetti L, Balconi M and Fazzi EM (2019) Visuospatial Attention and Saccadic Inhibitory Control in Children With Cerebral Palsy. Front. Hum. Neurosci. 13:392. doi: 10.3389/fnhum.2019.00392

Maitre, N. L., Key, A. P., Chorna, O. D., Slaughter, J. C., Matusz, P. J., Wallace, M. T., & Murray, M. M. (2017). The dual nature of early-life experience on somatosensory processing in the human infant brain. Current biology, 27(7), 1048–1054. doi: 10.1016/j.cub.2017.02.036

Marschik, P. B., Kwong, A. K., Silva, N., Olsen, J. E., Schulte-Rüther, M., Bölte, S., … & Spittle, A. J. (2023). Mobile solutions for clinical surveillance and evaluation in infancy—general movement apps. Journal of clinical medicine, 12(10), 3576. 10.3390/jcm12103576

Marschik, P. B., Pokorny, F. B., Peharz, R., Zhang, D., O’Muircheartaigh, J., Roeyers, H., & BEE-PRI study group. (2017). A novel way to measure and predict development: A heuristic approach to facilitate the early detection of neurodevelopmental disorders. Current neurology and neuroscience reports, 17(5), 43.

Matusz, P. J., Dikker, S., Huth, A. G., & Perrodin, C. (2019). Are we ready for real-world neuroscience?. Journal of cognitive neuroscience, 31(3), 327–338. 10.1162/jocn_e_01276#

Matusz, P. J., Key, A. P., Gogliotti, S., Pearson, J., Auld, M. L., Murray, M. M., & Maitre, N. L. (2018). Somatosensory plasticity in pediatric cerebral palsy following constraint-induced movement therapy. Neural plasticity, 2018(1), 1891978. 10.1155/2018/1891978

Mathiowetz, V., Federman, S., & Wiemer, D. (1985). Box and block test of manual dexterity: norms for 6–19 year olds. Canadian Journal of Occupational Therapy, 52(5), 241–245. 10.1177/000841748505200505

McIntyre, S., Goldsmith, S., Webb, A., Ehlinger, V., Hollung, S. J., McConnell, K., Arnaud, C., Smithers-Sheedy, H., Oskoui, M., Khandaker, G., & Himmelmann, K. (2022). Global prevalence of cerebral palsy: A systematic analysis. Developmental Medicine & Child Neurology, 64(12), 1494–1506. 10.1111/dmcn.15346

Mendoza-Sengco, P., Lee Chicoine, C., & Vargus-Adams, J. (2023). Early cerebral palsy detection and intervention. Paediatric Clinics of North America, 70(3), 385–398. 10.1016/j.pcl.2023.01.014

Miyake, A., Friedman, N. P., Emerson, M. J., Witzki, A. H., Howerter, A., & Wager, T. D. (2000). The unity and diversity of executive functions and their contributions to complex “frontal lobe” tasks: A latent variable analysis. Cognitive Psychology, 41(1), 49–100. 10.1006/cogp.1999.0734

Morgan, C., Fetters, L., Adde, L., Badawi, N., Bancale, A., Boyd, R. N., & Novak, I. (2021). Early intervention for children aged 0 to 2 years with or at high risk of cerebral palsy: international clinical practice guideline based on systematic reviews. JAMA pediatrics, 175(8), 846–858.

Mousavi, E., Akbarfahimi, N., Moein, S., & Vahedi, M. (2023). A study of the relationship between executive function and school function in children with cerebral palsy. Journal of Occupational Therapy, Schools, & Early Intervention, 16(2), 160–172.

Munn, Z., Pollock, D., Khalil, H., Alexander, L., Mclnerney, P., Godfrey, C. M., Peters, M., & Tricco, A. C. (2022). What are scoping reviews? Providing a formal definition of scoping reviews as a type of evidence synthesis. JBI Evidence Synthesis, 20(4), 950–952. 10.11124/JBIES-21-00483

Murray, M. M., Matusz, P. J., & Amedi, A. (2015). Neuroplasticity: unexpected consequences of early blindness. Current biology, 25, R998–R1001.

Novak, I., Morgan, C., Adde, L., Blackman, J., Boyd, R. N., Brunstrom-Hernandez, J., & Badawi, N. (2017). Early, accurate diagnosis and early intervention in cerebral palsy: advances in diagnosis and treatment. JAMA pediatrics, 171(9), 897–907.

Olsen, J., Marschik, P, & Spittle, A. (2018). Do fidgety general movements predict cerebral palsy and cognitive outcome in clinical follow-up of very preterm infants? Acta Paediatrica, 107(2), 361–362. Doi: 10.1111/apa.14126

Pant, S. W., Skovgaard, A. M., Ammitzbøll, J., Holstein, B. E., & Pedersen, T. P. (2022). Motor development problems in infancy predict mental disorders in childhood: a longitudinal cohort study. European Journal of Pediatrics, 181(7), 2655–2661. 10.1007/s00431-022-04462-3

Palisano, R. J., Hanna, S. E., Rosenbaum, P. L., Russell, D. J., Walter, S. D., Wood, E. P., & Galuppi, B. E. (2000). Validation of a model of gross motor function for children with cerebral palsy. Physical therapy, 80(10), 974–985. 10.1093/ptj/80.10.974

Paul, S., Nahar, A., Bhagawati, M., & Kunwar, A. J. (2022). A review on recent advances of cerebral palsy. Oxidative Medicine and Cellular Longevity, 2022(1), 2622310. 10.1155/2022/2622310

Piek, J. P., Dawson, L., Smith, L. M., & Gasson, N. (2008). The role of early fine and gross motor development on later motor and cognitive ability. Human movement science, 27(5), 668–681. doi: 10.1016/j.humov.2007.11.002

Pereira, A., Lopes, S., Magalhães, P., Sampaio, A., Chaleta, E., & Rosário, P. (2018). How executive functions are evaluated in children and adolescents with cerebral palsy? A systematic review. Frontiers in Psychology, 9, 21. 10.3389/fpsyg.2018.00021

Peters, M. D., Godfrey, C., McInerney, P., Munn, Z., Tricco, A. C., & Khalil, H. (2020). Chapter 11: Scoping reviews. In E. Aromataris & Z. Munn (Eds), JBI Manual for Evidence Synthesis (pp. 406–451). JBI. 10.46658/JBIMES-20-12.

Pirila, S., van der Meere, J. J., Rantanen, K., Jokiluoma, M., & Eriksson, K. (2011). Executive functions in youth with spastic cerebral palsy. Journal of child neurology, 26(7), 817–821. doi: 10.1177/0883073810392584.

Prechtl, H. F., Einspieler, C., Cioni, G., Bos, A. F., Ferrari, F., & Sontheimer, D. (1997). An early marker for neurological deficits after perinatal brain lesions. The Lancet, 349(9062), 1361–1363.

Rai, Y., Chaturvedi, S., Paliwal, V. K., Goyal, P., Chourasia, A., Rathore, R. K. S., & Gupta, R. K. (2013). DTI correlates of cognition in term children with spastic diplegic cerebral palsy. european journal of paediatric neurology, 17(3), 294–301. 10.1016/j.ejpn.2012.11.005.

Ramstad, K., Loge, J.H., Jahnsen, R. & Diseth, T.H. (2015). Self-reported mental health in youth with cerebral palsy and associations to recurrent musculoskeletal pain. Disabil. Rehabil., 37, 144–150. 10.3109/09638288.2014.913703

Reid, S. M., Ditchfield, M. R., Bracken, J., & Reddihough, D. S. (2015). Relationship between characteristics on magnetic resonance imaging and motor outcomes in children with cerebral palsy and white matter injury. Research in Developmental Disabilities, 45, 178–187. doi: 10.1016/j.ridd.2015.07.030.

Reilly, D. S., Woollacott, M. H., van Donkelaar, P., & Saavedra, S. (2008). The interaction between executive attention and postural control in dual-task conditions: Children with cerebral palsy. Archives of Physical Medicine and Rehabilitation, 89(5), 834–842. 10.1016/j.apmr.2007.10.023

Rethlefsen, S. A., Ryan, D. D., & Kay, R. M. (2010). Classification systems in cerebral palsy. The Orthopedic Clinics of North America, 41(4), 457–467. 10.1016/j.ocl.2010.06.005

Roid, G.H., Koch, C. (2017). Leiter-3: Nonverbal Cognitive and Neuropsychological Assessment. In: McCallum, R. (eds) Handbook of Nonverbal Assessment. Springer, Cham. 10.1007/978-3-319-50604-3_8

Romine, C. B., Lee, D., Wolfe, M. E., Homack, S., George, C., & Riccio, C. A. (2004). Wisconsin Card Sorting Test with children: a meta-analytic study of sensitivity and specificity. Archives of Clinical Neuropsychology, 19(8), 1027–1041. 10.1016/j.acn.2003.12.009

Rosenbaum, P., Paneth, N., Leviton, A., Goldstein, M., & Bax, M. (2006). The definition and classification of cerebral palsy. Developmental Medicine & Child Neurology, 49(109), 1–43. 10.1111/j.1469-8749.2007.00001.x

Salavati, S., Einspieler, C, Vagelli, G, Zhang, D, Pansy, J, Burgerhof, J. G. M., Marschik, P. B., Bos, A. F. (2017). The association between the early motor repertoire and language development in term children born after normal pregnancy. Early Human Development, 111(1), 30–35. Doi: 10.1016/j.earlhumdev.2017.05.006

Sergeant, J. (2000). The cognitive-energetic model: an empirical approach to attention-deficit hyperactivity disorder. Neuroscience & Biobehavioral Reviews, 24(1), 7–12. doi: 10.1016/S0149-7634(99)00060-3.

Shimko, G. A., & James, K. H. (2025). The relationship between motor development and ADHD: A critical review and future directions. Behavioral Sciences, 15, 576.

Sköld, A., Hermansson, L.N., Krumlinde-Sundholm, L., Eliasson, A.C. (2011). Development and evidence of validity for the Children’s Hand-use Experience Questionnaire (CHEQ). Developmental Medicine & Child Neurology, 53,436–42.

Smith, L. B., & Thelen, E. (2003). Development as a dynamic system. Trends in cognitive sciences, 7(8), 343–348. doi: 10.1016/S1364-6613(03)00156-6.

Simon-Martinez, C., Antoniou, M. P., Bouthour, W., Bavelier, D., Levi, D., Backus, B. T., & Matusz, P. J. (2023). Stereoptic serious games as a visual rehabilitation tool for individuals with a residual amblyopia (AMBER trial): a protocol for a crossover randomized controlled trial. BMC ophthalmology, 23, 220.

Sohlberg, M. M., & Mateer, C. A. (2001). Improving attention and managing attentional problems. Annals of the New York Academy of Sciences, 931(1), 359–375. 10.1111/j.1749-6632.2001.tb05790.x

Soriano, J. U., & Hustad, K. C. (2021). Speech-language profile groups in school aged children with cerebral palsy: Nonverbal cognition, receptive language, speech intelligibility, and motor function. Developmental Neurorehabilitation, 24(2), 118–129. 10.1080/17518423.2020.1858360

Spittle, A. J., Marschik, P. B., Adde, L., Badawi, N., Byrne, R., Bos, A. F., & Passmore, E. (2025). Towards universal early screening for cerebral palsy: a roadmap for automated General Movements Assessment. EClinicalMedicine, 86.

Spittle, A. J., & Morgan, C. (2025). Early intervention for children with cerebral palsy. In Cerebral Palsy: From Childhood to Adulthood (pp. 197–205). Cham: Springer Nature Switzerland.

Stadskleiv, K., Jahnsen, R., Andersen, G. L., & von Tetzchner, S. (2018). Neuropsychological profiles of children with cerebral palsy. Developmental Neurorehabilitation, 21(2), 108–120. 10.1080/17518423.2017.1282054

Surkar, S. M., Edelbrock, C., Stergiou, N., Berger, S., & Harbourne, R. (2015). Sitting postural control affects the development of focused attention in children with cerebral palsy. Paediatric Physical Therapy: The Official Publication of the Section on Paediatric s of the American Physical Therapy Association, 27(1), 16–22. 10.1097/PEP.0000000000000097

Te Velde, A., & Morgan, C. (2022). Gross Motor Function Measure (GMFM-66 & GMFM-88) User’s Manual, Book Review. doi: 10.1097/PEP.0000000000000858.

Thelen, E., & Smith, L. B. (1994). A dynamic systems approach to the development of cognition and action. The MIT press.

Tricco A.C., Lillie, E., Zarin, W., et al. (2018). PRISMA Extension for Scoping Reviews (PRISMA-ScR): Checklist and Explanation. Ann Intern Med., 169, 467–473. doi: 10.7326/M18-0850

Van der Fels, I. M. J., Te Wierike, S. C. M., Hartman, E., Elferink-Gemser, M. T., Smith, J., & Visscher, C. (2015). The relationship between motor skills and cognitive skills in 4–16 year old typically developing children: A systematic review. Journal of Science and Medicine in Sport, 18(6), 697–703. 10.1016/j.jsams.2014.09.007

Van Eck, M., Dallmeijer, A. J., Voorman, J. M., & Becher, J. G. (2009). Longitudinal study of motor performance and its relation to motor capacity in children with cerebral palsy. Developmental Medicine & Child Neurology, 51(4), 303–310. 10.1111/j.1469-8749.2008.03263.x

Van Rooijen, M., Verhoeven, L., Smits, D.-W., Ketelaar, M., Becher, J. G., & Steenbergen, B. (2012). Arithmetic performance of children with cerebral palsy: The influence of cognitive and motor factors. Research in Developmental Disabilities, 33(2), 530–537. 10.1016/j.ridd.2011.10.020

Van Rooijen, M., Verhoeven, L., & Steenbergen, B. (2016). Working memory and fine motor skills predict early numeracy performance of children with cerebral palsy. Child Neuropsychology, 22(6), 735–747. Embase. 10.1080/09297049.2015.1046426

Vitrikas, K., Dalton, H., & Breish, D. (2020). Cerebral Palsy: An Overview. American Family Physician, 101(4), 213–220. https://www.aafp.org/pubs/afp/issues/2020/0215/p213.html

Wallace, S. S., Barak, G., Truong, G., & Parker, M. W. (2022). Hierarchy of evidence within the medical literature. Hospital Paediatric s, 12(8), 745–750. 10.1542/hpeds.2022-006690

Wang, T.-N., Liang, K.-J., Howe, T.-H., Chen, H.-L., Huang, C.-W., & Wu, C.-T. (2020). Spatial attention disregard in children with hemiplegic cerebral palsy. The American Journal of Occupational TherapyfZ: Official Publication of the American Occupational Therapy Association, 74(2), 7402205090p1-7402205090p9. Medline. 10.5014/ajot.2020.038851

Wechsler, D. (2003). Wechsler Intelligence Scale for Children–Fourth Edition (WISC-IV): Administration and scoring manual. The Psychological Corporation.

Wechsler, D. (1997). Wechsler Adult Intelligence Scale–Third Edition (WAIS-III): Administration and scoring manual. The Psychological Corporation.

Wechsler, D. (1991). Wechsler Intelligence Scale for Children—Third Edition (WISC-III): Manual. The Psychological Corporation.

Wechsler, D., & Naglieri, J.A. (2006). Wechsler nonverbal scale of ability (WNV). Pearson.

Wotherspoon, J., Whittingham, K., Sheffield, J., & Boyd, R. (2023). Executive function, attention and autism symptomatology in school-aged children with cerebral palsy. Journal of Developmental and Physical Disabilities, 36, 187–202. 10.1007/s10882-023-09905-9

Zhou, Y., & Tolmie, A. (2024). Associations between gross and fine motor skills, physical activity, executive function, and academic achievement: longitudinal findings from the UK millennium cohort study. Brain Sciences, 14(2), 121. doi 10.3390/brainsci14020121

Zhu, F., Zhu, X., Bi, X., Kuang, D., Liu, B., Zhou, J., … & Ren, Y. (2023). Comparative effectiveness of various physical exercise interventions on executive functions and related symptoms in children and adolescents with attention deficit hyperactivity disorder: A systematic review and network meta-analysis. Frontiers in Public Health, 11, 1133727. 10.3389/fpubh.2023.1133727.

